# Landscape of SARS-CoV-2 genomic surveillance, public availability extent of genomic data, and epidemic shaped by variants: a global descriptive study

**DOI:** 10.1101/2021.09.06.21263152

**Authors:** Zhiyuan Chen, Andrew S. Azman, Xinhua Chen, Junyi Zou, Yuyang Tian, Ruijia Sun, Xiangyanyu Xu, Yani Wu, Wanying Lu, Shijia Ge, Zeyao Zhao, Juan Yang, Daniel T. Leung, Daryl B. Domman, Hongjie Yu

**Author notes:** Corresponding authors: Hongjie Yu, School of Public Health, Fudan University, Shanghai 200032, China. **Disclaimer:** The views expressed are those of the authors and do not necessarily represent the institutions with which the authors are affiliated.

## Abstract

**Background:** Genomic surveillance has shaped our understanding of SARS-CoV-2 variants, which have proliferated globally in 2021. Characterizing global genomic surveillance, sequencing coverage, the extent of publicly available genomic data coupled with traditional epidemiologic data can provide evidence to inform SARS-CoV-2 surveillance and control strategies.

**Methods:** We collected country-specific data on SARS-CoV-2 genomic surveillance, sequencing capabilities, public genomic data, and aggregated publicly available variant data. We divided countries into three levels of genomic surveillance and sequencing availability based on predefined criteria. We downloaded the merged and deduplicated SARS-CoV-2 sequences from multiple public repositories, and used different proxies to estimate the sequencing coverage and public availability extent of genomic data, in addition to describing the global dissemination of variants.

**Findings:** Since the start of 2021, the COVID-19 global epidemic clearly featured increasing circulation of Alpha, which was rapidly replaced by the Delta variant starting around May 2021 and reaching a global prevalence of 96.6% at the end of July 2021. SARS-CoV-2 genomic surveillance and sequencing availability varied markedly across countries, with 63 countries performing routine genomic surveillance and 79 countries with high availability of SARS-CoV-2 sequencing. Less than 3.5% of confirmed SARS-CoV-2 infections were sequenced globally since September 2020, with the lowest sequencing coverage in the WHO regions of Eastern Mediterranean, South East Asia, and Africa. Across different variants, 28-52% of countries with explicit reporting on variants shared less than half of their variant sequences in public repositories. More than 60% of demographic and 95% of clinical data were absent in GISAID metadata accompanying sequences.

**Interpretation:** Our findings indicated an urgent need to expand sequencing capacity of virus isolates, enhance the sharing of sequences, the standardization of metadata files, and supportive networks for countries with no sequencing capability.

**Research in context:** 

**Evidence before this study:** On September 3, 2021, we searched PubMed for articles in any language published after January 1, 2020, using the following search terms: (“COVID-19” OR “SARS-CoV-2”) AND (“Global” OR “Region”) AND (“genomic surveillance” OR “sequencing” OR “spread”). Among 43 papers identified, few papers discussed the global diversity in genomic surveillance, sequencing, public availability of genomic data, as well as the global spread of SARS-CoV-2 variants. A paper from Furuse employed the publicly GISAID data to evaluate the SARS-CoV-2 sequencing effort by country from the perspectives of “fraction”, “timeliness”, and “openness”. Another viewpoint paper by Case Western Reserve University’s team discussed the impediments of genomic surveillance in several countries during the COVID-19 pandemic. The paper as reported by Campbell and colleagues used the GISAID data to present the global spread and estimated transmissibility of recently emerged SARS-CoV-2 variants. We also found several studies that reported the country-level genomic surveillance and spread of variants. To our knowledge, no research has quantitatively depicted the global SARS-CoV-2 genomic surveillance, sequencing ability, and public availability extent of genomic data.

**Added value of this study:** This study collected country-specific data on SARS-CoV-2 genomic surveillance, sequencing capabilities, public genomic data, and aggregated publicly available variant data as of 20 August 2021. We found that genomic surveillance strategies and sequencing availability is globally diverse. Less than 3.5% of confirmed SARS-CoV-2 infections were sequenced globally since September 2020. Our analysis of publicly deposited SARS-CoV-2 sequences and officially reported number of variants implied that the public availability extent of genomic data is low in some countries, and more than 60% of demographic and 95% of clinical data were absent in GISAID metadata accompanying sequences. We also described the pandemic dynamics shaped by VOCs.

**Implications of all the available evidence:** Our study provides a landscape for global sequencing coverage and public availability extent of sequences, as well as the evidence for rapid spread of SRAS-CoV-2 variants. The pervasive spread of Alpha and Delta variants further highlights the threat of SARS-CoV-2 mutations despite the availability of vaccines in many countries. It raised an urgent need to do more work on defining the ideal sampling schemes for different purposes (e.g., identifying new variants) with an additional call to share these data in public repositories to allow for further rapid scientific discovery.

## Introduction

Following the first pandemic wave of coronavirus disease 2019 (COVID-19), the emergence and dissemination of SARS-CoV-2 variants have resulted in new waves of infections across the globe in 2021. Some SARS-CoV-2 variants disappeared immediately, while others characterized by several key mutations adapted well, enabling their rapid spread^1^. WHO has designated four Variants of Concern (VOCs) associated with increased transmissibility and various extents of immune escape^2–4^, namely the Alpha, Beta, Gamma, and Delta variants, first detected in the UK, South Africa, Brazil, and India, respectively^5^. Specifically, the Delta variant is highly transmissible, with an estimated transmissibility increase of 40-60% compared with Alpha variant^6–9^, while the Beta variant has been shown to have the highest reduction in neutralization activity whether from natural infection or vaccination^10^, both reflected in lower vaccine efficacy or effectiveness^11–13^.

The identification and classification of SARS-CoV-2 variants mainly relied on partial or whole genome sequencing, although PCR assays have been used to identify specific features relatively unique in specific variants, like spike gene target failure (SGTF)^14^. Since the first SARS-CoV-2 sequence was published in January 2020^15^, the unprecedented rate of genome data generation was far greater than any other pathogen^16^, with 3.1 million genomes deposited in Global Initiative on Sharing All Influenza Data (GISAID) through August 2021^17^. Genomic data has been vital to the early detection of mutations and monitoring of virus evolution, as well as evaluating the degree of similarities between circulating variants with vaccine strains, especially since the availability of SARS-CoV-2 vaccines^18^.

Several studies have employed genomic data to examine the evolution and associated spread of dominant variants in one country or one region, raising a claim of the rapidity of local transmission of SARS-CoV-2 variants and urgency of genomic surveillance^6,19–21^. However, the lack of representation of genomic data from low and middle-income countries (LMICs) was most concerning in these studies ^6,19–21^. Indeed, the impact of genome data is dependent on their quality, and the reliability and accuracy of such data may influence the global community’s ability to track the spread of variants in a timely manner.

In this study, we aimed to investigate the global diversity of SARS-CoV-2 genomic surveillance and the global sequencing coverage of confirmed cases and public availability extent of genomes. In addition, we sought to map the global identification and spread of SARS-CoV-2 variants. This data can provide evidence to better inform policy on SARS-CoV-2 surveillance.

## Methods

### Data sources and collection

Through extracting country-specific data from multiple publicly available sources, we built three datasets of genomic surveillance, deposited genomic data in publicly repositories, and official aggregated genomic data as of 20 August 2021.

#### Dataset of genomic surveillance

Each country’s genomic surveillance strategy and sequencing availability was gathered from searches of the websites of regional WHO, the country’s Ministry of Health, CDC, local academic partners, and official news, supplemented by a literature search (appendix). Data extracted included the overall surveillance strategy, sequencing availability, target population, sampling method, diagnostic criteria, and sequenced volume. Given that the surveillance strategy and density may change with time, we only gathered information on the most recent surveillance strategy (as of 20 August 2021).

#### Dataset of SARS-CoV-2 sequences in publicly repositories

SARS-CoV-2 sequences along with related metadata file were downloaded from an online coronavirus analysis platform from the National Genomics Data Center (NGDC)^22^, where has merged and deduplicated sequences that deposited in GISAID^23^, GenBank^24^, National Genomics Data Center^25^, National Microbiology Data Center^26^, and China National GeneBank^27^. Detailed process of integration and deduplication was previously reported^28^. An acknowledgement table for those contributing to this work is available in the Appendix.

#### Official aggregated dataset

To gain additional insights regarding the public availability (sharing) extent of genomic data of SARS-CoV-2 variants, we extracted country-specific, variant-specific, and time-specific aggregated data on the number of SARS-CoV-2 variant cases from official websites, using the same sources as above, except for the literature source. The search was done by either directly locating to the official website for each country or indirectly searching in search engines (Google, Bing, Baidu) by using the terms “variant” and country name. To supplement the aggregated data of variants that we collected, we also downloaded the aggregated data with a valid denominator (namely, the number of isolates sequenced is reasonable) from the European Surveillance System (TESSy). The aggregated dataset included country name, date of report or collection, new or cumulative numbers of different SARS-CoV-2 variant cases and total sequenced cases, as well as the new number of confirmed cases. COVID-19 epidemic data were derived from WHO^29^. Considering that the diagnostic criteria of SARS-CoV-2 variants varies in different countries, the general principle for collecting aggregated data was to give priority to the results based on whole and partial genome sequencing instead of those based on a PCR assay.

For countries noted by WHO as having had VOCs identified, but no data in publicly repositories, we collected the information about when VOC/VOCs were first detected from country’s Ministry of Health and official media news. The search of media news was also done in search engine queries (Google, Bing, Baidu) using combined terms “first” and “variant” and country name.

All data were entered into a structured database by a trained team (co-authors). All recorded data were cross-checked by coauthors. Details of data sources and data completeness are shown in Appendix (Table S1-S2, S4-S5).

### Data analysis

We used the variant naming system proposed by WHO, where four VOCs and five Variants of Interest (VOIs, included Eta, Iota, Kappa, Lambda, and Mu) had been designated as of 2 September 2021^5^. Our study focused on analyses on four VOCs in the 194 Member States of WHO. We did not integrate data from the overseas territories into that country’s data. Given most countries weekly released aggregated data of variant, most of our analyses were performed on a weekly basis.

To characterize the global diversity of SARS-CoV-2 genomic surveillance, we classified the surveillance strategy of each country into three categories: 1) routine genomic surveillance; 2) limited routine genomic surveillance; and 3) no routine genomic surveillance. Routine genomic surveillance was defined as conducting nationwide genomic sequencing, coupled with at least 150 specimens per week or 10% of all samples sequenced^30^. The classification of most African countries was from a pre-defined version from African CDC^31^, while other countries were subsequently defined according to the definition criteria (Table S3). As we could not identify information on the surveillance strategy for some countries through public sources, we also classified three extra categories according to the public availability/ability of genomic sequencing: 1) high availability; 2) moderate availability; and 3) low availability (Table S3).

The process of data cleaning in publicly repositories and aggregated dataset is shown in the Appendix. For data that was reported in aggregate, when the date of sample collection was not available, we assumed a fixed three-week lag^32^ from sample collection to reporting unless other country-specific information was available to inform this extrapolation.

The sequencing coverage was inferred by using the percent of cumulative positives sequenced as a proxy, defined by the ratio of the number of isolates sequenced to the number of confirmed cases in the same unit of time. Given that not all sequences will be uploaded to genomic repositories, we analysed the public availability extent of genomic data by comparing the cumulative number of variants between public repositories and official aggregated datasets. Since the Alpha variant had a characteristic deletion of amino acids 69-70 in the SGTF that can be detected via a widely used PCR assay^33^, we performed this analysis across VOCs.

We plotted the earliest time when the first VOC specimen was identified in each country. The earliest identification was defined by the earliest sampling time of sequences deposited in publicly repositories. If a VOC was identified by WHO but no corresponding sequence in publicly repositories for one country, we used the date collected from other sources. The sequences with sampling date earlier than the earliest sample identified in the United Kingdom (for Alpha), South Africa (for Beta), Brazil (for Gamma) and India (for Delta), respectively, were not used in analyses.

We also described the global and regional prevalence trend of variants. The prevalence of variant was defined as the proportion of the variant number to the sequencing number in the same period. When multiple data sources were available for one country, the most abundant dataset was chosen. For example, if there was publicly available genomic data as well as an official aggregated dataset, the priority was given to the one with highest reported sequenced numbers in a specific week.

Statistical significance was tested using the Chi-square test for comparing ratios of two groups, and differences were considered statistically significant at P-value < 0.05. All statistical analyses and visualizations were done using R (version 4.0.2).

## Results

We classified genomic surveillance strategies for 105 countries, including 72.3% (34/47) of WHO-defined Africa Region countries, 58.5% (31/53) of European Region countries, 61.9% (13/21) of countries in the Eastern Mediterranean Region, 31.4% (11/35) of countries in Americas Region, 37.0% (10/27) of countries in the Western Pacific Region, and 54.5% (6/11) of countries in the South East Asia Region (Table S4). We downloaded a total of 3.1 million deduplicated SARS-CoV-2 sequence samples from publicly repositories corresponding to samples collected between 1 December 2019 to 20 August 2021 in 163 countries. Additionally, we collected official aggregated data of variants from 55 countries, and extra data for the first identification of COVID-19 variants from 33 countries.

### The global diversity of SARS-CoV-2 genomic surveillance strategies and sequencing availability

We observed marked geographical heterogeneity in genomic surveillance of SARS-CoV-2 across countries. Globally, a total of 60.0% (63) countries had performed routine genomic surveillance, 26.7% (28) countries implemented limited routine genomic surveillance, 13.3% (14) countries had no routine genomic surveillance with the remaining countries (89) having no data on genomic surveillance strategy identified (Figure 1A). Surveillance diversity across various countries was also reflected in the context of target populations, sampling method, sequenced proportion, and diagnostic criteria (Table S4). Specifically, 32 countries randomly selected or used all confirmed cases with sufficient quality for sequencing, and 18 countries adopted PCR assay to screen or confirm variants. From the regional perspective, limited or no routine genomic surveillance was common in the Eastern Mediterranean (84.6%, 11/13) and Africa (61.8%, 21/34), followed by the South East Asia (50.0%, 3/6), Americas (36.4%, 4/11), Western Pacific (20.0%, 2/10), and Europe (3.2%, 1/31) (Figure 1A). However, among the 167 Member States with data accessible, the availability of SARS-CoV-2 sequences was high in 79 countries, moderate in 80 countries, and low in 8 countries (Figure 1B).

**Fig 1.**
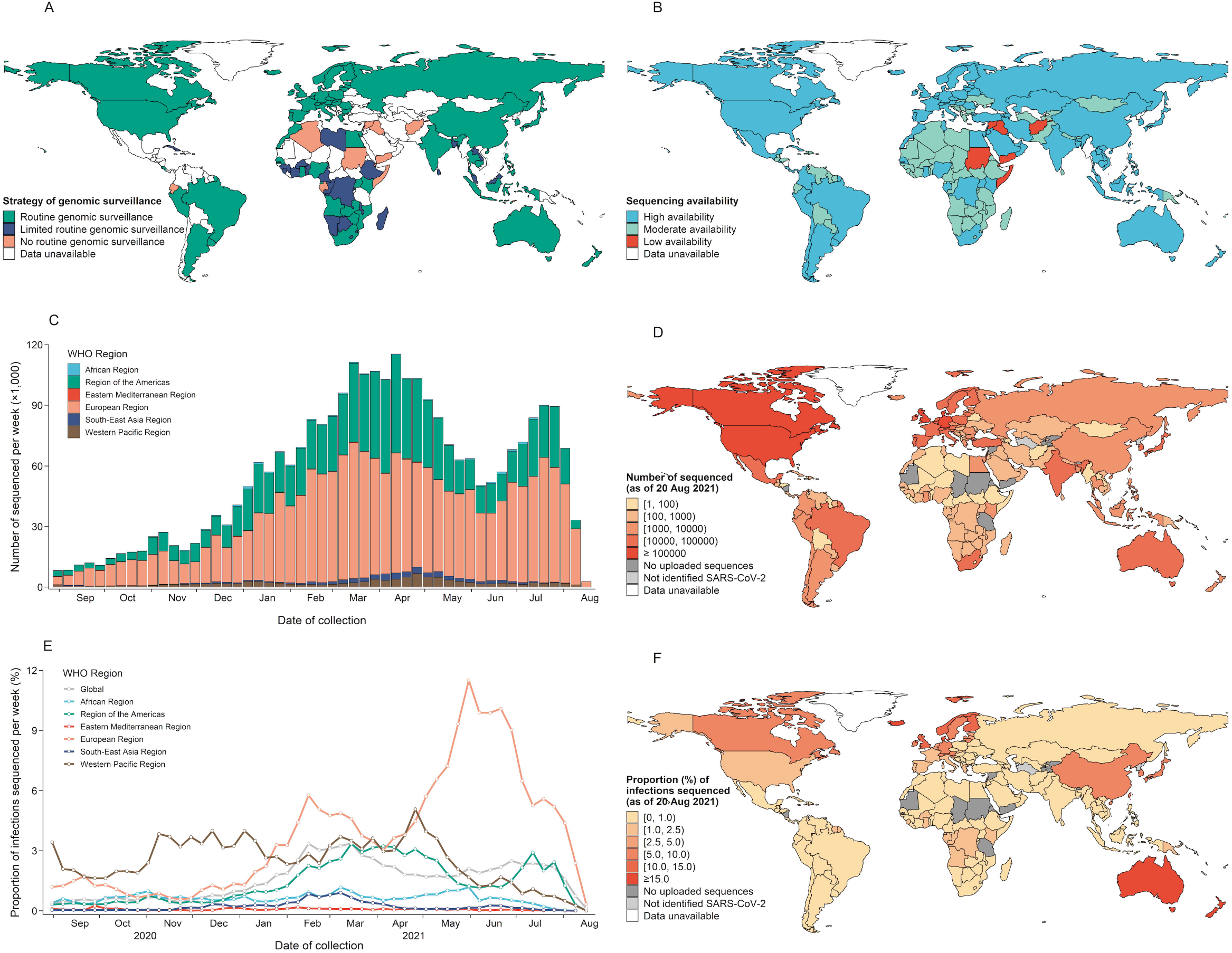
Global SARS-CoV-2 genomic surveillance, sequencing availability, and publicly deposited genomic data. (A) The global distribution of three strategies of SARS-CoV-2 genomic surveillance. (B) The global availability of SARS-CoV-2 sequencing, countries with a high level of availability represent the ability to perform in-country SARS-CoV-2 sequencing alone. (C) The weekly number of publicly deposited SARS-CoV-2 genomic data by region. (D) Cumulative number of publicly deposited SARS-CoV-2 genomic data by countries as of 20 August 2021. (E) The weekly proportion of infections sequenced by region. (F) Cumulative proportion of infections sequenced by countries as of 20 August 2021, defined as the proportion of cumulative isolates sequences to the cumulative confirmed cases. The number of sequences for the most recent weeks might be incomplete due to a time delay between collecting specimens and sequencing submission. Data unavailable, includes those locations that not belonged to 194 Member States or had not applicable data.

### Sequencing coverage of SARS-CoV-2 confirmed cases

The global volume of genomic data gradually increased over time until April 2021 (Figure 1C). The European (59.4%) and Americas (33.2%) Regions uploaded the most sequences to public repositories, with marked intra-region heterogeneity across countries, with a range from 3 (Saint Kitts and Nevis) to 847,000 (United States) as of 20 August 2021 (Figure 1D).

Since September 2020, no more than 3.5% of global confirmed SARS-CoV-2 infections were sequenced. In any week, the Africa, South East Asia and Eastern Mediterranean Regions (Figure 1E) sequenced no more than 1.2% of cases. At the end of May 2021(start of Delta widely spreading), Europe had the highest sequenced proportion of 9.3%, followed by Western Pacific (2.1%), Americas (1.4%), Africa (1.0%), South-East Asia (0.1%), and Eastern Mediterranean (0.07%) (Figure 1E). At the country-level, higher rates of sequencing were observed in Iceland, New Zealand, Australia, Denmark, Luxembourg, Finland, United Kingdom, and Norway, all of which had at least 10% reported infections sequenced as of 20 August 2021. In addition, almost all countries in the Africa and Eastern Mediterranean had sequenced less than 2.5% of confirmed infections, except for Gambia (6.6%) and Mauritius (3.5%) (Figure 1F).

### Public availability extent of SARS-CoV-2 variant sequences

The public availability extent of SARS-CoV-2 genomic data varied across variants and countries. Overall, among countries with aggregated data on the number of variant infections, less than half of sequences of Alpha, Beta, Gamma, and Delta were publicly available in 45.3% (24/53), 27.9% (12/43), 33.3% (11/33), and 52.3% (23/44) countries, respectively (Figure 2). However, the result for Alpha might be influenced by SGTF detected via PCR. The public availability extent of Delta variants across countries ranged from 0.0% (Cyprus, Hungary, Iceland, Laos) to 92.9% (Denmark). At the country level, low sharing proportions (less than 50%) across all VOCs was found in several countries, including Austria, Cyprus, Greece, Hungary, Iceland, Philippines, Thailand and Senegal. For example, the sharing proportion of Alpha, Beta, and Delta in Thailand was 5.2%, 13.6%, and 2.0%, respectively, which indicated more than 85.0% genomic data of variants were not uploaded.

**Fig 2.**
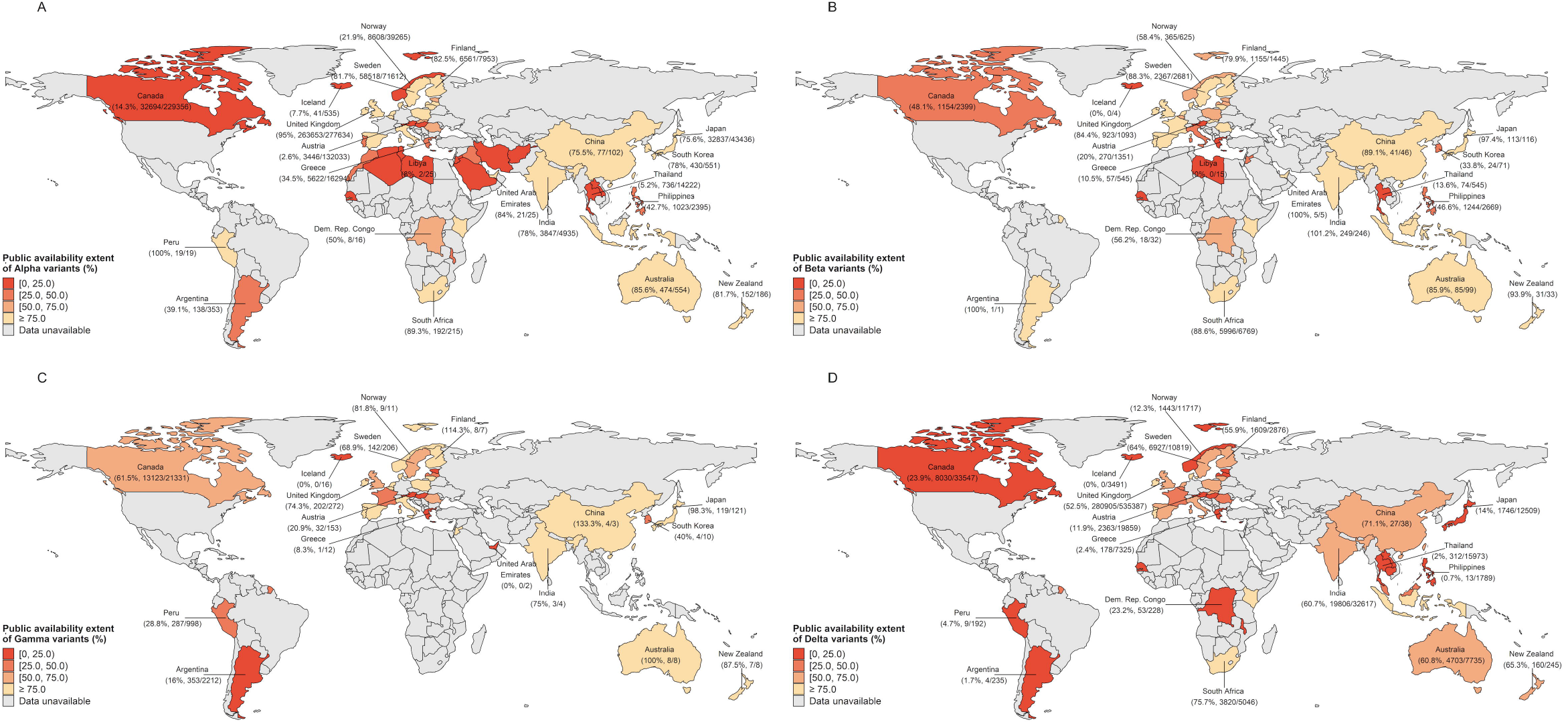
The public availability extent of SARS-CoV-2 genomic data to publicly repositories by country and the variants of concern. The public availability extent was defined as the ratio of the cumulative number of variants in publicly repositories to the official reported number of variants within the same period. The ratio of exceed 100% in some countries might be due to the delay in the official report of the sequencing results or the incomplete official reporting system. In view of the availability of official data, the cumulative number of variants in different countries corresponds to different time periods, with detailed information in Table S6. Sequence without date of specimen collection in publicly repositories is not included in our analysis. The variant data for China only includes cases reported by mainland China. The officially reported number of Alpha variants might contain those confirmed by the PCR screen assay. The values beneath the country names show numbers of cumulative variant as of one specific week: variants in publicly repositories/official reported variants.

Moreover, incomplete metadata attached to GISAID sequences was common globally, with about two thirds of sequences missing demographic information (age and sex), and more than 95% of that missing clinical information (e.g., symptom history, clinical outcome, and vaccination status) (Table S5). We found that high-income regions tend to have lower information completeness (P < 0.0001, Chi-square test), especially in the European Region, where less than 25.0% and 3.0% of sequences had demographic and clinical information, respectively. Besides, 92.8% sequences were reported by subnational geographies, with a low proportion in Western Pacific (52.0%) and Eastern Mediterranean (76.9%).

### Earliest identification of SARS-CoV-2 variants across regions

Alpha was first identified in the Europe, then in Americas and South-East Asia in September-October 2020, followed by spread to Eastern Mediterranean, Africa, and Western Pacific in November 2020 (Figure 3A). The earliest publicly available sequenced Beta infection was sampled in Africa in May 2020, and subsequently identified in other regions (Figure 3B). Gamma variant detection remained spatially constrained after it was first identified in Brazil (Figure 3C). After the first identification of Delta in October 2020 in South-East Asia, global spread occurred after January 2021 (Figure 3D).

**Fig 3.**
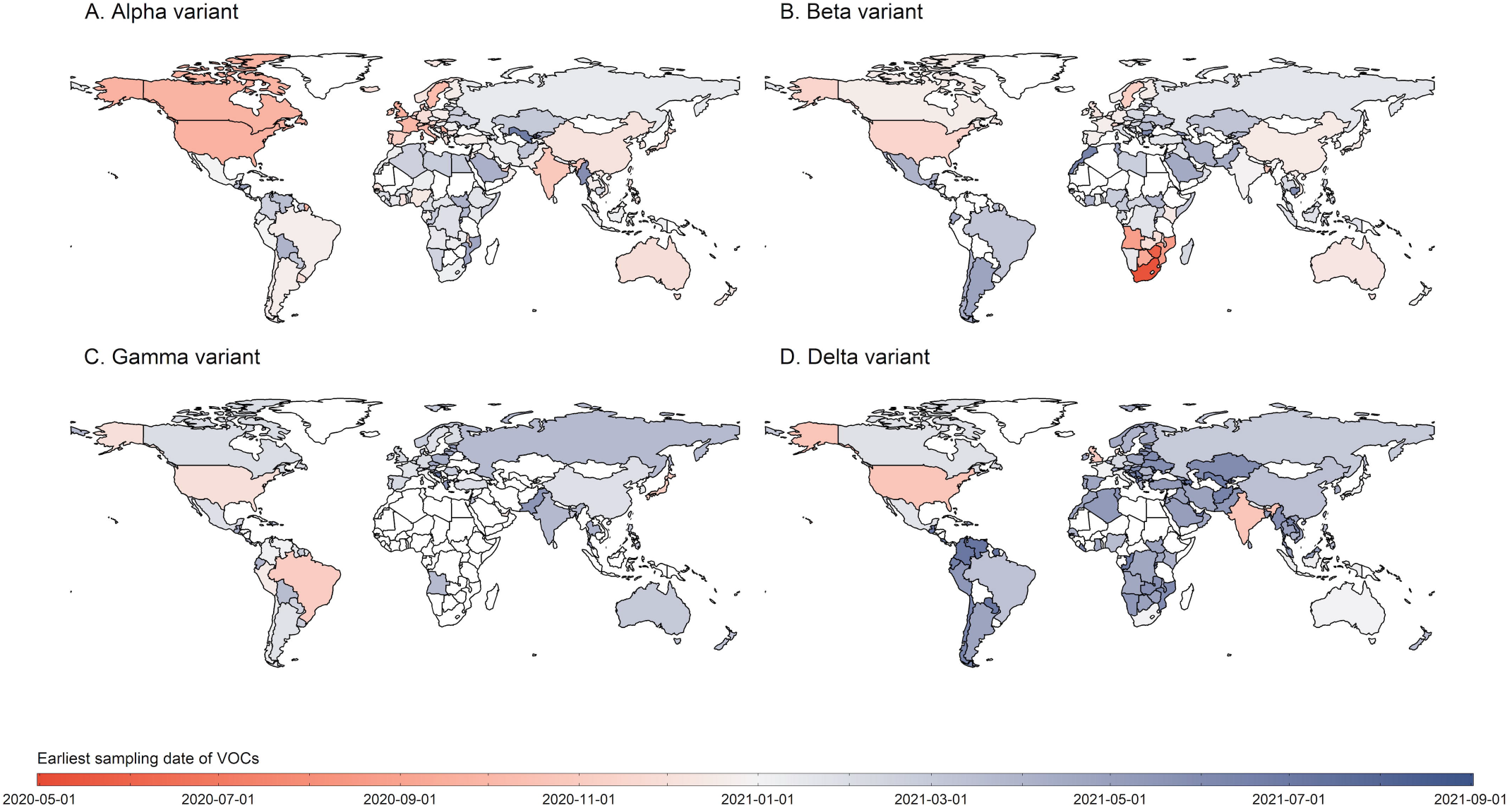
The earliest identification of Alpha, Beta, Gamma, and Delta variant in each country. If information about the earliest sampling date was unavailable but that of the earliest reporting date was available, we extrapolated the sampling date by using a fixed three-week lag from sample collection to reporting. Countries with darker red indicate earlier samples, and with darker blue refers to later samples. The white areas represent countries with variants unreported or data unavailable.

### Global and regional spread of SARS-CoV-2 variants

The number of new VOC cases dramatically increased until April 2021, with a peak weekly value of about 100,000 VOC cases sequenced in which most of them were Alpha variants (Figure 5A). Subsequently, another peak of weekly new VOC case occurred in July 2021, but with a large amount of Delta variants. The number of VOC cases may be an underestimate for the most recent weeks due to a collection-to-report time delay. Notably, this increase was also accompanied by the increase in the volume of new sequenced cases and new COVID-19 confirmed cases.

The global prevalence of reference (non-variant) strains fell into a low level of 0.6% in the period of Jul-Aug 2021, compared with 13.4% of that in 2020 (Figure 4). Globally, the COVID-19 pandemic was driven by the circulation of Alpha at the start of 2021, with an average prevalence of 50.6% in the first quarter of 2021. Alpha variants continued to outcompete other strains in the second quarter of 2021, accounting for 60.0% of the contemporary lineages (Figure 4). However, the rapid global rise of the Delta variant began in May 2021, reaching a global prevalence of nearly 96.6% at the end of July 2021 (Figure 5B). In contrast, Beta and Gamma variants remained at low prevalence (Figure 4). Additionally, the shifting of predominant variants from Alpha to Delta first occurred in South-East Asia where the proportion of Delta exceeded 60.0% in April 2021 (Figure 5J).

**Fig 4.**
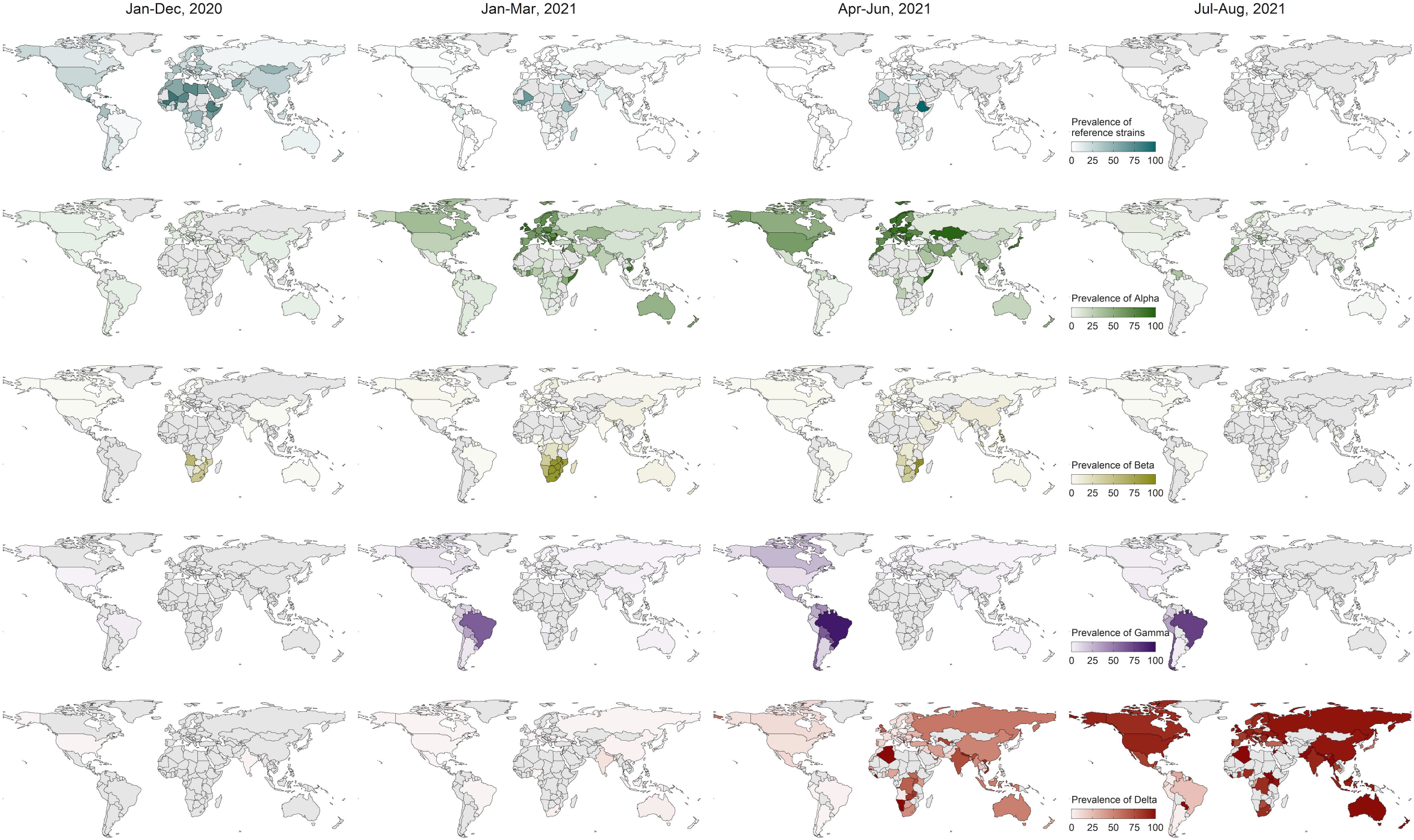
The prevalence and temporal dynamics of reference strains and four SARS-CoV-2 VOCs by country. The date presented in the top refers to the range of date of specimen collection. The prevalence was defined as the proportion of the strain number (reference strains or variants) to the total number of sequences generated in the same unit of time. Reference strains includes lineage A, A.1, B, and B.1; the sub-lineages of four VOCs are aggregated with the parent lineages. The grey areas represent countries with no COVID-19 epidemic, or no performing sequencing, or no uploading genomic data to publicly repositories.

**Fig 5.**
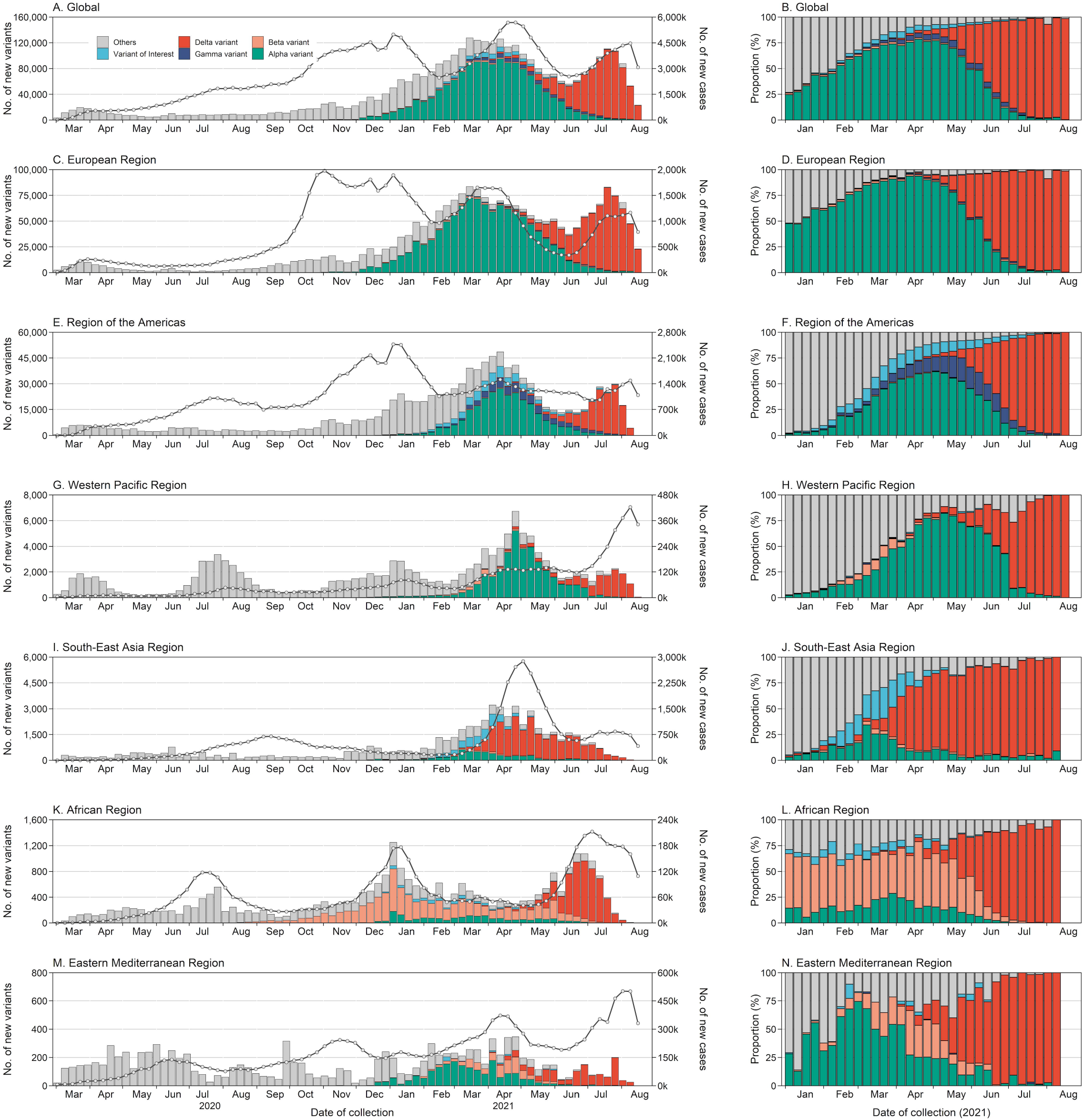
The number and proportion of SARS-CoV-2 variants by region and time. The line and point in the left figure correspond to the y-axis on the right. The sub-lineages of four VOCs are aggregated with the parent lineages; designated Variants of Interest (VOIs) included lineage B.1.525, B.1.526, B.1.617.1, C.37, B.1.621 and their sub-lineages; other lineages included reference strains and other variants. Data used here are derived from the publicly repositories and aggregated dataset, with priority given to the one with highest sequenced numbers in a specific week.

## Discussion

Our study characterized the global diversity of genomic surveillance strategies and sequencing availability, sequenced coverage of SARS-CoV-2 cases, public availability extent of variant sequences, as well as current epidemic situation of SARS-CoV-2 variants. We found that genomic surveillance strategies were globally heterogenous, with limited or no routine surveillance among many countries in the Africa and Eastern Mediterranean Regions. Our analysis of publicly deposited SARS-CoV-2 sequences implied that the sequenced coverage is low in most countries, with a low proportion of VOCs sequences shared to public repositories. The pervasive spread of Alpha and Delta variants further highlights the threat of SARS-CoV-2 mutations despite the availability of vaccines in many countries. Overall, we describe the pandemic dynamics shaped by VOCs and call for more work on defining the ideal sampling schemes for different purposes (e.g., identifying new variants) with an additional call to share these data in public repositories to allow for further rapid scientific discovery.

The diversity of SARS-CoV-2 genomic surveillance between countries is associated with country-specific priorities (e.g., surveillance objectives, targeted monitoring, or event-/risk-based sequencing) and available resources. ECDC recommends population-based and/or targeted sampling strategies (e.g., imported cases, cluster cases, and potential vaccine escapers) for genomic surveillance, which could provide a more representative estimate of the relative prevalence of variants. Notably, several countries, many of which are classified as low- or lower middle-income countries by the World Bank, lack genomic surveillance data, likely due to limitations in infrastructure capacity and resources. However, even some countries classified as high-income, have suffered from a slow and inconsistent process of adopting genomics-based surveillance^34^. Establishment of reference laboratories and networks to provide sequencing services for countries without established sequencing capacity may enable improved detection and tracking of emerging variants worldwide.

The detection of most variants relies on the full-length or partial genomic sequencing, but the sequences only become available for the global community when the laboratories have established sequencing capacity, willing to share, and legally allowed to upload them. The discrepancies in sharing was observed in each region, which confirmed that some countries are sequencing but are not uploading. However, our study observed a sharing extent of exceed 100% exists in some countries, likely due to delays in the official reporting of sequencing results, or the incomplete official reporting system. The timely sharing of those enables to adequately contextualize local data when looking at introductions and examine transmission routes, as well as to look for sites of repeated mutations that can guide laboratory work on characterizing those mutations effects on therapeutics and vaccine efficacy. The underlying reasons why some countries didn’t share might be related with the distrust for publicly repositories in the concept of data security.

We found relatively low completeness of demographic and clinical characteristics in metadata accompanying uploaded sequences. Our analysis revealed that high-income countries frequently did not share demographic information. A possible reason for this is that these regions may having more restrictive data privacy/laws preventing/discouraging the release of this information. Genomic data coupled with those additional data can maximize the utility of genomic sequences in rapid scientific discovery during this pandemic, which are valuable for in-depth epidemiological analyses to characterize risk factors, clinical severity, and other public health risk of variants^35–37^. Therefore, it’s vital to optimize the sharing of information in a secure and trusted channel in the context of protect patient anonymity and in accordance of local regulations^38^. In addition, decreasing the lag between sample collection to deposition of these sequences^39^, including the timely sharing and standardizing of metadata ^18,30,35^, may facilitate the design and development of treatment and prevention strategies by policy-makers^40^.

An important role of genomic surveillance is to investigate the spread and dynamics of SARS-CoV-2 variants. Amidst the emergence of different variants, the current dominance of the Delta variant suggests that it may possess higher fitness than other variants, which might be associated with a combination of higher virus load, and shorter incubation period and serial intervals^41–43^. The decrease in real-world vaccine effectiveness against Beta variants^11^ and increasing breakthrough cases with Delta variants^44^ underlines the importance of determining the local or regional patterns of variant spread, including the need to develop new or modified vaccines to achieve adequate protection^45^.

Our results should be interpreted in view of several limitations. First, the lack of data from some countries limited our global mapping. The data completeness and quality could be impacted by key steps in the surveillance or reporting, including differences in diagnostic criteria, under-reporting, delayed reporting, and reporting methods. The inconsistent diagnostic criteria of variants might cause sampling bias, especially when adopting PCR assay to detect Alpha variant owing to its non-specificity^46^. We did an extensive search to collect multi-source data and chose the aggregated data with a priority to sequencing results rather than PCR-screening results. Second, the analysis of global and national spread could be biased as data from public repositories or aggregated dataset are not always representative of the variants circulating in the regions, especially for the regions with relatively limited sequencing capacity or with investigating outbreak-based events. Therefore, the global prevalence of variants may be biased due to the uneven sequencing across the regions. Indeed, it’s difficult to obtain a truly representative and random sample, and how to understand these biases will become important^36^. Lastly, the detailed demographical, epidemiological and clinical information about variant cases cannot be accessed, which limited our further epidemiological analysis about variant spread.

In conclusion, our study provides a landscape for genomic surveillance, the global coverage of sequencing and public availability of sequences, as well as the evidence for rapid spread of SARS-CoV-2 variants. Our findings suggest that global SARS-CoV-2 genomic surveillance strategies and capacity are diverse, and may be limited in some regions, especially in the context of the global spread and dominance of variants of concern. The gap still exists in sequencing availability and magnitude, therefore international efforts are needed to address some genomic bottleneck, such as lack of tracking representative samples, lack of sequencing capacity, strict regulations about data sharing, and lack of funding ^46,47^.

## Data Availability

All datasets generated and analysed are available in the Article and Appendix. Any additional information is available from the lead contact upon request.

## Contributors

H.Y. designed and supervised the study. Z.C., J.Z., Y.T., R.S, X.X., Y.W., W.L., S.G., and Z.Z. collected data. Z.C., J.Z., Y.T., R.S, and X.X. prepared the tables. Z.C. analysed data, prepared the figures, and wrote the first draft of the manuscript. A.S.A., X.C., J.Y., D.T.L., D.B.D, and H.Y. interpreted the results and revised the content critically. All authors contributed to review and revision and approved the final manuscript as submitted and agree to be accountable for all aspects of the work.

## Acknowledgments

We thank all the authors contributed to generating and sharing sequences to GISAID, GenBank, National Genomics Data Center, National Microbiology Data Center, and China National GeneBank. We thank all the authors contributed to generating and sharing aggregated data to TESSy in ECDC. We thank Dr.Lina Ma, Dr.Yiming Bao, and their team from China National Center for Bioinformation for building 2019nCoVR dataset. We thank Junbo Chen, Xiaowei Deng from Fudan University. This study was funded by Key Program of the National Natural Science Foundation of China (grant no. 82130093 to HJY) and the US National Institutes of Health (R01 AI135115 to DTL and ASA; KL2TR001448 to DD).

