## Appendix for "Landscape of SARS-CoV-2 genomic surveillance, public availability extent of genomic data, and epidemic shaped by variants: a global descriptive study"

### Methods

#### Nomenclature of SARS-CoV-2 variant

The dynamic SARS-CoV-2 nomenclature system from Phylogenetic Assignment of Named Global Outbreak Lineages (PANGOLIN) adopted a phylogenetic framework to identify new lineages[1], which was previously most employed. On 31 May 2021, WHO announced a new naming system that using letters of the Greek Alphabet (i.e., Alpha for B.1.1.7) for easy-to-easy and coherent application[2], and hereafter would be used in our study. Based on comparative assessment on phenotypic impact of SARS-CoV-2 variants, some variants with significant signals would be classified into variants of concern (VOCs) and variants of interest (VOIs).

#### Literature search for genomic surveillance

Data of genomic surveillance was supplemented by literature search. We searched PubMed and Europe PMC for peer-reviewed and preprint studies that characterized the country-level strategy of SARS-CoV-2 genomic surveillance from January 1, 2020 to Aug 20, 2021. The search was done with the following terms: “SARS-CoV-2”, “COVID-19”, “sequencing”, and “genomic surveillance”. Articles that published in English and contained information about genomic surveillance and sequencing capability will be included. The data from literature will entered the structured dataset.

#### Data cleaning in publicly repositories and aggregated dataset

For sequences in publicly repositories, we removed sequences of the non-human host and non-assignment of PANGO lineage. After that, all assigned PANGO lineages will be classified into seven categories (reference strains, Alpha, Beta, Gamma, Delta, VOI, and other variants) based on the designation of WHO. The reference strains included lineage A, B, with additional lineage A.1 and B.1 that harbored D614G mutation in spike protein. The sub-lineages of four VOCs are aggregated with the parent lineages, and designated VOIs included Eta, Iota, Kappa, Lambda, and Mu based on the WHO categories. Sequences were also removed if date of collection was incomplete (only year) or sampled before 1 December 2019. When explicit dates were not provided but with a month provided, we selected the mid-month as the date of sampling.

For the analyses of official aggregated data, cases of alpha variant included cases with or without E484K mutation site, and delta variant included lineage B.1.617.2 and sub-lineages of AY. For the date of reporting in aggregated dataset, we employed a fixed three-week lag to extrapolate date of collection[3], unless the lay information was known for this country.

#### Classification of genomic surveillance and sequencing availability

WHO recommends an ideal number of 150 representative specimens per week or 10% of samples to be sequenced for countries establishing sentinel surveillance systems[4], which hence was regarded as the definition of routine genomic surveillance in our study. The Limited routine genomic surveillance was defined as countries that sequenced at least 15 nationwide specimens per week or ≥1% of all positive samples. If countries sequenced less than 15 nationwide specimens per week and <1% of all positive samples, we regarded as no routine genomic surveillance. In addition, we also outlined the global sequencing availability, which has been classified into three categories: high availability, moderate availability, and low availability.

High availability was defined as the one that can collect viral isolates from clinical samples and conduct in-country genomic sequencing. Countries that using regional sequencing networks or need to ship samples to external labs outside countries were placed in the category of moderate availability (Table S3). The regional networks in Africa contained several reference laboratories to provide services to countries in their sub-regions[5], therefore the countries where the reference laboratories were located were defined as "high availability" and those countries served by the reference laboratories were defined as "moderate availability". If countries had no sequencing capability and with little supportive sequencing services from the external labs, we placed them in the category of low availability.

### Supplementary Tables

#### Table S1. Data sources for aggregated dataset on SARS-CoV-2 variants.

| Country name | Main sources |
|---|---|
| **EUR** | |
| Switzerland | Federal Office of Public Health (https://www.covid19.admin.ch/en/epidemiologic/virus-variants?geo=CH&time=total) |
| Sweden | The Swedish Public Health Agency (https://www.folkhalsomyndigheten.se/smittskydd-beredskap/utbrott/aktuella-utbrott/covid-19/statistik-och-analyser/sars-cov-2-virusvarianter-av-sarskild-betydelse/) |
| United Kingdom | Public Health England (https://www.gov.uk/government/publications/covid-19-variants-genomically-confirmed-case-numbers) |
| Spain | Ministry of Health, Consumption and Social Welfare (https://www.mscbs.gob.es/profesionales/saludPublica/ccayes/alertasActual/nCov/documentos) |
| Luxembourg | National Health Laboratory (https://lns.lu/departement/microbiologie/revilux/) |
| Netherlands | National Institute for Public Health and the Environment (https://www.rivm.nl/en/coronavirus-covid-19/virus-sars-cov-2/variants) |
| Norway | The National Institute of Public Health (https://www.fhi.no/en/publ/2020/weekly-reports-for-coronavirus-og-covid-19/) |
| Ireland | Health Protection Surveillance center (https://www.hpsc.ie/a-z/respiratory/coronavirus/novelcoronavirus/surveillance/summaryofcovid-19virusvariantsinireland/Virus%20Variants%20report.pdf) |
| Bulgaria | National Center for Infectious and Parasitic Diseases (https://www.ncipd.org/index.php?option=com_k2&view=item&id=546:ncov-012020&lang=bg) |
| Denmark | Danish Covid-19 Genome Consortium (https://www.covid19genomics.dk/statistics) |
| Finland | Department of Health and Welfare (https://thl.fi/fi/web/infektiotaudit-ja-rokotukset/ajankohtaista/ajankohtaista-koronaviruksesta-covid-19/tilannekatsaus-koronaviruksesta) |
| Austria | Austria Agency for Food Safety (https://www.ages.at/themen/krankheitserreger/coronavirus/sars-cov-2-varianten-in-oesterreich/) |
| Germany | The Robert Koch Institute (https://www.rki.de/DE/Content/InfAZ/N/Neuartiges_Coronavirus/DESH) |
| France | Public health of France (https://www.santepubliquefrance.fr/maladies-et-traumatismes/maladies-et-infections-respiratoires/infection-a-coronavirus/documents/bulletin-national) |

| | |
|---|---|
| Other European countries | European CDC (TESSy) (https://www.ecdc.europa.eu/en/publications-data/data-virus-variants-covid-19-eueea) |
| **AMR** | |
| Canada | CTV New.ca's variant tracker (https://www.ctvnews.ca/health/coronavirus/tracking-variants-of-the-novel-coronavirus-in-canada-1.5296141) |
| United States | US. CDC (1. https://www.cdc.gov/mmwr/volumes/70/wr/pdfs/mm7023a3-H.pdf; 2. https://covid.cdc.gov/covid-data-tracker/#variant-proportions) |
| Argentina | 1. Ministry of Health (https://www.argentina.gob.ar/salud/coronavirus-COVID-19/informacion-epidemiologica/julio-2021) 2. Argentine Interinstitutional SARS-CoV-2 Genomic Project (1. http://pais.qb.fcen.uba.ar/files/reportes; 2. https://www.argentina.gob.ar/sites/default/files/2021/06/ reporte_ndeg23_vigilancia_activa_de_variantes_extendido_de_sars-cov2_07-06-2021.pdf) |
| Brazil | Ministry of Health (https://www.gov.br/saude/pt-br/media/pdf/2021) |
| Peru | Ministry of Health (https://web.ins.gob.pe/es/covid19/secuenciamiento-sars-cov2) |
| **SEAR** | |
| Bangladesh | International Centre for Diarrhoeal Disease Research (https://www.icddrb.org/news-and-events/news?id=874&task=view) |
| India | Institute of Genomics and Integrative Biology (http://clingen.igib.res.in/covid19genomes/) |
| Indonesia | Health Research and Development Agency (http://www.litbang.kemkes.go.id/assets/2021/06/Update_VoC-COVID-19-20-juni-2021.pdf) |
| Thailand | Department of Medical Sciences Ministry of Health (https://www3.dmsc.moph.go.th/post-group/10) |
| **WPR** | |
| Australia | 1. The CDGN VOC Taskforce (https://www.cdgn.org.au/variants-of-concern) 2. Ministry of Health (https://www1.health.gov.au/internet/main/publishing.nsf /Content/1D03BCB527F40C8BCA258503000302EB/$File) |
| South Korea | Korea Disease Control and Prevention Agency (http://www.kdca.go.kr/board/board.es?mid=a30501000000&bid=0031&cg_code=C05) |
| Philippines | Department of health (https://doh.gov.ph/) |
| Malaysia | Ministry of Health |

| | |
|---|---|
| | (http://covid-19.moh.gov.my/semasa-kkm/2021) |
| New Zealand | Ministry of Health<br>(1. https://www.health.govt.nz/system/files/documents/pages;<br>2. https://www.health.govt.nz/our-work/diseases-and-conditions/<br>covid-19-novel-coronavirus/covid-19-resources-and-tools/covid-19-science-news) |
| Japan | Ministry of Health, Labour and Welfare & National institute of infectious diseases<br>(1. https://www.mhlw.go.jp/content/10900000/000812902.pdf<br>2.https://www.niid.go.jp/niid/ja/diseases/ka/corona-virus/2019-ncov/2484-idsc/<br>10279-covid19-40.html) |
| Laos | WHO Regional for the WPR<br>(https://www.who.int/laos/emergencies/covid-19-in-lao-pdr/situation-reports) |
| Cambodia | WHO Regional for the WPR<br>(https://www.who.int/cambodia/emergencies/covid-19-response-in-cambodia/situation-reports) |
| **AFR** | |
| African countries | Africa CDC<br>(https://africacdc.org/institutes/africa-pathogen-genomics-initiative/) |
| **EMR** | |
| Countries in the EMR | WHO Regional for the EMR<br>(http://www.emro.who.int/health-topics/corona-virus/situation-reports.html) |

**Table S2. Data sources for the first identification of SARS-CoV-2 variants.**

| Country name | Source type | Date type (variants) | Main sources |
|---|---|---|---|
| EUR | | | |
| Belarus | Media news | Date of report (Delta) | https://www.uniindia.com/belarus-reports-first-case-of-covid-19-delta-variant-ministry-of-health/world/news/2429337.html |
| Cyprus | Media news | Date of report (Beta) | http://www.xinhuanet.com/english/2021-05/20/c_139956862.htm |
| Estonia | Media news | Date of report (Gamma) | https://news.err.ee/1608226756/brazilian-indian-coronavirus-strains-recorded-in-estonia |
| Hungary | Media news | Date of report (Beta) | https://hungarytoday.hu/coronavirus-south-african-variant-hungary-muller/ |
| | | Date of report (Delta) | https://xpatloop.com/channels/2021/06/nine-delta-coronavirus-variant-cases-identified-in-hungary.html |
| Kazakhstan | Media news | Date of report (Beta) | https://astanatimes.com/2021/03/kazakh-capital-city-announces-new-restrictions-as-cases-surge/ |
| | | Date of report (Delta) | https://ca.movies.yahoo.com/kazakhstan-detects-delta-variant-central-090939082.html |
| Kyrgyzstan | Media news | Date of collection (Alpha) | http://med.kg/en/news/4380-four-covid-19-variants-are-circulating-in-kyrgyzstan.html |
| | | Date of collection (Beta) | http://med.kg/en/news/4380-four-covid-19-variants-are-circulating-in-kyrgyzstan.html |
| Ukraine | Media news | Date of report (Beta) | https://www.reuters.com/article/health-coronavirus-ukraine-idUSS8N2JP00X |
| | | Date of report (Delta) | https://www.reuters.com/business/healthcare-pharmaceuticals/ukraine-registers-first-cases-covid-19-delta-variant-2021-06-23/ |
| Uzbekistan | Media news | Date of collection (Alpha)* | https://for.kg/news-687161-en.html |
| | | Date of report (Delta) | https://akipress.com/news:659766:Uzbekistan_detects_Delta_variant_of_COVID-19/ |
| AMR | | | |
| Antigua and Barbuda | Media news | Date of collection (Beta) | https://caribbean.loopnews.com/nl/node/550384 |
| Bolivia | Media news | Date of report (Alpha) | https://brazilian.report/liveblog/2021/04/22/bolivia-british-coronavirus-variant/ |
| Cuba | Media news | Date of report (Alpha) | https://www.miamiherald.com/news/nation-world/world/americas/cuba/article248782480.html |

| | | | |
|---|---|---|---|
| | | Date of report (Beta) | |
| Guatemala | Media news | Date of report (Beta) | https://en.memesrandom.com/guatemala-suspects-the-presence-of-delta-variant-of-coronavirus-and-will-send-50-samples-for-analysis-in-a-laboratory-in-panama-prensa-libre/ |
| | | Date of report (Gamma) | |
| Panama | Media news | Date of report (Gamma) | https://www.cnn.com/world/live-news/coronavirus-pandemic-vaccine-updates-01-23-21/h_ce583284d449a8da7f427104f69c584e |
| Uruguay | Media news | Date of report (Alpha) | https://www.republica.com.uy/hallaron-la-variante-britanica-del-virus-sars-cov-2-en-uruguay-id810761/ |
| SEAR | | | |
| Maldives | Media news | Date of report (Alpha) | https://en.sun.mv/67274 |
| WPR | | | |
| Brunei [a] | Media news | Date of report (Alpha) | http://www.moh.gov.bn/Lists/Latest%20news/NewDispForm.aspx?ID=816 |
| Fiji | Media news | Date of report (Delta) | https://www.france24.com/en/live-news/20210427-fiji-fears-virus-tsunami-after-outbreak-found-to-be-indian-variant |
| Laos | Official data | Date of report (Alpha) | https://www.who.int/docs/default-source/wpro---documents/countries/lao-people's-democratic-republic/covid-19/covid_19_wco-moh_sitrep_29-20210428.pdf?sfvrsn=18ca45ab_5 |
| | Media news | Date of report (Delta) | https://www.thestar.com.my/aseanplus/aseanplus-news/2021/07/01/laos-records-first-cases-of-delta-variant |
| Vietnam [b] | Media news | Date of collection (Beta) | https://e.vnexpress.net/news/news/vietnam-detects-1st-case-of-south-african-coronavirus-variant-infection-4229589.html |
| AFR | | | |
| Algeria | Media news | Date of collection (Alpha) | https://www.aps.dz/sante-science-technologie/118228-covid-19-deux-cas-du-variant-britannique-decouverts-en-algerie |
| | | Date of report (Delta) | https://www.reuters.com/business/healthcare-pharmaceuticals/algeria-finds-first-cases-indian-coronavirus-variant-2021-05-03/ |
| Cape Verde | Media news | Date of report (Alpha) | https://inforpress.cv/covid-19-ministry-of-health-confirms-circulation-of-the-english-variant-in-cabo-verde/ |
| Liberia | Official data | Date of report (Alpha) | https://africacdc.org/download/outbreak-brief-59-coronavirus-disease-2019-covid-19-pandemic/ |
| Mozambique | Media news | Date of report (Delta) | https://allafrica.com/stories/202106220975.html |

| | | | |
|---|---|---|---|
| Namibia | Media news | Date of report (Beta) | http://www.xinhuanet.com/english/africa/2021-03/11/c_139801765.htm |
| Seychelles [a] | Media news | Date of collection (Beta) | http://www.seychellesnewsagency.com/articles/14680/COVID+testing+shows+that+South+African+variant+is+present+in+Seychelles |
| Zimbabwe [b] | Media news | Date of collection (Delta) | https://www.reuters.com/world/africa/zimbabwe-reports-first-cases-coronavirus-variant-india-2021-05-19/ |
| EMR | | | |
| Afghanistan | Official data | Date of report (Alpha, Delta) | 1.http://www.emro.who.int/images/stories/coronavirus/documents/covid_19_bi_weekly_sitrep_7.pdf?ua=1 2.http://www.emro.who.int/images/stories/coronavirus/revised-15.pdf?ua=1 |
| Lebanon | Media news | Date of report (Delta) | https://www.thenationalnews.com/mena/lebanon/2021/07/02/lebanon-records-first-three-cases-of-covid-19-delta-variant/ |
| Libya | Official data | Date of report (Beta) | http://www.emro.who.int/images/stories/coronavirus/12.pdf?ua=1 |
| Pakistan | Official data | Date of report (Gamma) | http://www.emro.who.int/images/stories/coronavirus/12.pdf?ua=1 |
| Saudi Arabia | Official data | Date of report (Delta) | http://www.emro.who.int/images/stories/coronavirus/12.pdf?ua=1 |
| Tunisia | Media news | Date of report (Delta) | https://allafrica.com/stories/202106280113.html |
| United Arab Emirates | Official data | Date of report (Gamma) | http://www.emro.who.int/images/stories/coronavirus/12.pdf?ua=1 |
| | Media news | Date of report (Delta) | https://www.thenationalnews.com/uae/health/highly-contagious-delta-variant-accounts-for-one-in-three-uae-cases-1.1249954 |

[a] Exact date was not reported, and with only month information available. Here we used the 15th of each month to replace it.

[b] Arrival time from other countries. Here we assumed that the sample will be immediately collected after landing.

**Table S3. Definition of different genomic surveillance strategy and sequencing availability.**

| **Surveillance strategy** | **Definition** |
|---|---|
| Genomic surveillance strategy [a] | |
| Routine genomic surveillance | Conduct nationwide genomic sequencing, coupled with at least 150 specimens per week or 10% of all samples sequenced[4,6]. |
| Limited routine genomic surveillance | Conduct nationwide genomic sequencing, coupled with at least 15 specimens per week or 1% of all samples sequenced. |
| No routine genomic surveillance | Conduct genomic sequencing, coupled with less than 15 specimens per week and 1% of all samples sequenced. |
| Availability of genomic sequencing | |
| High availability | Be able to collect viral isolates from clinical samples and conduct in-country genomic sequencing. |
| Moderate availability | Be able to collect viral isolates from clinical samples, but the process of genomic sequencing needs extra supports from external sequencing labs, including the following scenarios: 1) samples need to ship to the regional reference labs or high-incoming countries for sequencing, such as Bolivia[7]; 2) purchase the commercial kits for the detect SARS-CoV-2 variants or get a donation of that, such as Ukraine[8]; 3) establish the sequencing laboratory with the support from others during the pandemic of COVID-19, such as Mongolia[9]. |
| Low availability | Lack of sequencing capability and have little supportive sequencing services from the external labs. |

[a] The category of genomic surveillance strategy was developed on the basis of a category from Africa CDC Institute of Pathogen Genomics[10].

**Table S4. Country-specific SARS-CoV-2 genomic surveillance strategy.**

| Country | Surveillance strategy | Sequencing availability | Target population | Sampling method | Sequenced volume | Reporting frequency | Diagnostic criteria | Source |
|---|---|---|---|---|---|---|---|---|
| **European Region (EUR)** | | | | | | | | |
| Austria | Routine genomic surveillance since April 2021 | High availability | 1) Positive RT-PCR cases<br>2) Visitors | Randomly selected (sentinel system) | 150 or more samples sequenced per week; at least 10% of samples sequenced | Weekly | Sequencing or PCR | https://www.ages.at/themen/krankheitserreger/coronavirus/sars-cov-2-varianten-in-oesterreich/ |
| Armenia | - | High availability | SARS-CoV-2 positive samples | - | - | - | Whole genome sequencing | 1.https://armenpress.am/eng/news/1047911.html<br>2.Avetyan D, et al. medRxiv 2021: 2021.06.19.21259172.[11] |
| Azerbaijan | - | High availability | Persons who arrived from abroad | - | - | Regularly | Sequencing | https://jam-news.net/delta-and-alpha-strains-of-covid-19-detected-in-azerbaijan/ |
| Belarus | - | High availability | 1) People who arrived from abroad<br>2) Confirmed COVID-19 cases | - | - | Continuously | Genotyping, sequencing | 1)https://www.belarus.by/en/press-center/press-release/uk-variant-of-coronavirus-reaches-belarus_i_126790.html<br>2) https://apa.az/en/xeber/cis-countries-news/belarus-reports-first-case-of-covid-19-delta-variant-ministry-of-health-says-352358 |
| Kazakhstan | - | High availability | Positive RT-PCR cases | Sampling in some regions and cities | 414 samples screened as of July 1 | - | Whole genome sequencing, PCR | https://astanatimes.com/2021/07/covid-19-delta-variant-confirmed-in-kazakhstan/ |
| Kyrgyzstan, Ukraine, Tajikistan, and Albania | - | Moderate availability | - | - | - | - | - | 1. https://www.reuters.com/business/healthcare-pharmaceuticals/uk-track-covid-19-variants-with-genomic-sequencing-across-world-2021-07-07/ |

| Country | Surveillance strategy | Sequencing availability | Target population | Sampling method | Sequenced volume | Reporting frequency | Diagnostic criteria | Source |
|---|---|---|---|---|---|---|---|---|
| | | | | | | | | 2. http://www.med.kg/en/news/4592-germany-provided-reagents-for-identifying-covid-19-variants-circulating-in-kyrgyzstan.html 3.https://www.who.int/publications/m/item/weekly-operational-update-on-covid-19---4-august-2021 |
| Belgium | Routine genomic surveillance | High availability | Positive RT-PCR cases | - | 150 or more samples sequenced per week | Weekly | Sequencing | https://www.ecdc.europa.eu/en/publications-data/data-virus-variants-covid-19-eueea |
| Bulgaria | Routine genomic surveillance | Moderate availability | Positive RT-PCR cases | Sampling in multiple districts | 150 or more samples sequenced per week | Weekly or daily | Sequencing | 1)https://www.ncipd.org/index.php?option=com_k2&view=item&id=546:ncov-012020&lang=bg 2)https://www.ecdc.europa.eu/en/publications-data/data-virus-variants-covid-19-eueea 3.https://euobserver.com/coronavirus/151238 |
| Switzerland | Routine genomic surveillance | High availability | Confirmed cases | Randomly selected | 150 or more samples sequenced per week; 5-9% of all confirmed cases per week | Weekly | Whole genome Sequencing | 1)https://cevo-public.github.io/Quantification-of-the-spread-of-a-SARS-CoV-2-variant/ 2)https://www.swissinfo.ch/eng/switzerland-shifts-focus-to-tracking-indian-covid-19-variant/ 3)https://www.covid19.admin.ch/en/epidemiologic/virus-variants?detTime=total&detGeo=CH 4) Chen et al. medRxiv 2021: 2021.03.05.21252520. [12] |
| Cyprus | Limited routine genomic surveillance | Moderate availability | Positive RT-PCR cases | - | 150 or more samples sequenced per week | Weekly | Sequencing | 1.https://www.ecdc.europa.eu/en/publications-data/data-virus-variants-covid-19-eueea 2.https://euobserver.com/coronavirus/151238 |

| **Country** | **Surveillance strategy** | **Sequencing availability** | **Target population** | **Sampling method** | **Sequenced volume** | **Reporting frequency** | **Diagnostic criteria** | **Source** |
|---|---|---|---|---|---|---|---|---|
| Czech Republic | Routine genomic surveillance | High availability | Positive RT-PCR cases | - | 150 or more samples sequenced per week | Weekly | Sequencing | https://www.ecdc.europa.eu/en/publications-data/data-virus-variants-covid-19-eueea |
| Germany | Routine genomic surveillance | High availability | 1) SARS-CoV-2 cases with a Ct value ≤ 25<br>2) Sequencing if a VOC is suspected | Randomly selected | 150 or more samples sequenced per week; 5% (10% if weekly cases < 70,000) of samples sequenced | Weekly | Partial and whole genome Sequencing, PCR | 1) https://www.rki.de/DE/Content/InfAZ/N/Neuartiges_Coronavirus/DESH/Handlungsanleitung_Labore.html;jsessionid=E1E13A97BA3E4222033D30%E2%80%A6<br>2) https://www.bundesgesundheitsministerium.de/fileadmin/Dateien/3_Downloads/C/Coronavirus/Verordnungen/CorSurV_Ref_mit_Begruendungsteil.pdf<br>3) https://www.bundesgesundheitsministerium.de/presse/pressemitteilungen/2021/1-quartal/coronavirus-surveillanceverordnung.html<br>4) https://www.rki.de/DE/Content/InfAZ/N/Neuartiges_Coronavirus/DESH/Bericht_VOC_2021-06-16.pdf?__blob=publicationFile |
| Denmark | Routine genomic surveillance Since March 2020 | High availability | SARS-CoV-2 positive samples from both pillars of testing | Sequenced almost all cases if can produce a genome of sufficient quality | 1000 or more samples sequenced per week; at least 80% of samples sequenced | Weekly | Sequencing | https://www.covid19genomics.dk/statistics |
| Spain | Routine genomic surveillance | High availability | 1) Community patients<br>2) Individuals with breakthrough infection | Randomly selected (nationwide) | 1000 or more samples sequenced per week | Weekly | Whole genome sequencing, PCR | https://www.mscbs.gob.es/profesionales/saludPublica/ccayes/alertasActual/nCov/situacionActual.htm |

| Country | Surveillance strategy | Sequencing availability | Target population | Sampling method | Sequenced volume | Reporting frequency | Diagnostic criteria | Source |
|---|---|---|---|---|---|---|---|---|
| Estonia | Routine genomic surveillance | High availability | Positive RT-PCR cases | - | 150 or more samples sequenced per week | Weekly | Sequencing | https://www.ecdc.europa.eu/en/publications-data/data-virus-variants-covid-19-eueea |
| Finland | Routine genomic surveillance | High availability | 1) COVID-19 cases<br>2) Positive samples taken from imported cases and their contacts<br>3) Samples taken from abnormal clusters of infections | - | 200–300 samples sequenced per week; about 20% of samples sequenced | Weekly | whole genome sequencing, whole S gene sequencing, or partial S gene sequencing | 1) https://thl.fi/fi/web/infektiotaudit-ja-rokotukset/taudit-ja-torjunta/taudit-ja-taudinaiheuttajat-a-o/koronavirus-covid-19/koronaviruksen-covid-19-laboratoriotutkimukset/naytteiden-lahettaminen-koronasekvensointiin<br>2) https://thl.fi/en/web/infectious-diseases-and-vaccinations/what-s-new/coronavirus-covid-19-latest-updates/transmission-and-protection-coronavirus/coronavirus-variants<br>3) https://www2.helsinki.fi/en/researchgroups/covid-19/virus-sequencing<br>4) https://thl.fi/fi/web/infektiotaudit-ja-rokotukset/ajankohtaista/ajankohtaista-koronaviruksesta-covid-19/tilannekatsaus-koronaviruksesta<br>5) https://thl.fi/en/web/infectious-diseases-and-vaccinations/what-s-new/coronavirus-covid-19-latest-updates/trans |
| France | Routine genomic surveillance | High availability | Positive RT-PCR samples | Randomly selected (Flash surveys) | 150 or more samples sequenced per week | Bi-weekly | Partial and whole genome Sequencing | 1)https://www.santepubliquefrance.fr/dossiers/coronavirus-covid-19/coronavirus-circulation-des-variants-du-sars-cov-2#block-338801<br>2) Quéromès G, et al. Emerg Microbes Infect 2021; 10(1): 167-77.[13] |

| Country | Surveillance strategy | Sequencing availability | Target population | Sampling method | Sequenced volume | Reporting frequency | Diagnostic criteria | Source |
|---|---|---|---|---|---|---|---|---|
| United Kingdom | Routine genomic surveillance | High availability | 1) COVID-19 patients in hospitals<br>2) Individuals from general community and care home<br>3) Imported cases | Randomly selected (nearly from each region) | Over 50% (depends on infection rate) of samples sequenced | Weekly | Sequencing and genotyping | 1)https://www.gov.uk/guidance/surge-testing-for-new-coronavirus-covid-19-variants#history<br>2)https://www.cogconsortium.uk/how-do-we-collect-and-sequence-sars-cov-2-samples/<br>3) Grubaugh ND, et al. Cell 2021; 184(5): 1127-32.[14] |
| Greece | Routine genomic surveillance | Moderate availability | Positive RT-PCR cases | - | 150 or more samples sequenced per week | Weekly | Sequencing | 1.https://www.ecdc.europa.eu/en/publications-data/data-virus-variants-covid-19-eueea<br>2.https://euobserver.com/coronavirus/151238 |
| Croatia | Routine genomic surveillance | Moderate availability | Positive RT-PCR cases | - | 150 or more samples sequenced per week; at least 10% of samples sequenced recently | Weekly | Sequencing | 1.https://www.ecdc.europa.eu/en/publications-data/data-virus-variants-covid-19-eueea<br>2. Rokić F, et al. Arch Virol. 2021;166(6):1735-1739.[15]<br>3. https://zdravstvo.gov.hr/vijesti/u-odlukama-o-popustanju-mjera-razmotrit-ce-se-porast-slucajeva/5438 |
| Hungary | Routine genomic surveillance | High availability | Positive RT-PCR cases | - | At least 10% of samples sequenced recently | Weekly | Sequencing | https://www.ecdc.europa.eu/en/publications-data/data-virus-variants-covid-19-eueea |
| Ireland | Routine genomic surveillance | High availability | 1) Incoming travelers<br>2) Complex clusters<br>3) Potential vaccine escape<br>4) Persons under investigation for VOC<br>5) Probable and confirmed cases | - | 1000 or more samples sequenced per week; 10% of samples sequenced (since week 51) | Weekly | Whole genome sequencing, PCR | 1)https://www.hpsc.ie/a-z/respiratory/coronavirus/novelcoronavirus/surveillance/summaryofcovid-19virusvariantsinireland/<br>2)https://www.hpsc.ie/a-z/respiratory/coronavirus/novelcoronavirus/sars-cov-2variantsofconcern |

| Country | Surveillance strategy | Sequencing availability | Target population | Sampling method | Sequenced volume | Reporting frequency | Diagnostic criteria | Source |
|---|---|---|---|---|---|---|---|---|
| Iceland | Routine genomic surveillance | High availability | Positive RT-PCR cases | - | At least 10% of samples sequenced | Weekly | Sequencing | https://www.ecdc.europa.eu/en/publications-data/data-virus-variants-covid-19-eueea |
| Israel | - | High availability | 1) Laboratory confirmed COVID-19 patients, including pregnant women<br>2) Positive individuals who will return from specific destinations | Randomly selected | - | Irregularly | Sequencing | 1)https://www.gov.il/en/Departments/news/<br>2) Munitz A, et al. Cell Rep Med 2021; 2(5): 100264.[16]<br>3)https://www.gov.il/en/departments/news/01022021-02 |
| Italy | Routine genomic surveillance | High availability | Positive RT-PCR cases | Voluntary samples | 150 or more samples sequenced per week | Weekly | Sequencing | 1)https://www.ecdc.europa.eu/en/publications-data/data-virus-variants-covid-19-eueea<br>2) Di Giallonardo F, et al. Viruses 2021; 13(5).[17] |
| Lithuania | Routine genomic surveillance | High availability | Confirmed cases | - | 150 or more samples sequenced per week; at least 10% of samples sequenced | Weekly | Sequencing | 1)https://www.ecdc.europa.eu/en/publications-data/data-virus-variants-covid-19-eueea<br>2)https://www.delfi.lt/en/politics/first-case-of-covid-19-delta-variant-detected-in-lithuania.d?id=87496673 |
| Luxembourg | Routine genomic surveillance | High availability | 1) Positive cases who are hospitalized and from Airport testing program<br>2) Outbreaks and identified clusters<br>3) Reinfections and post-vaccination-infections | Randomly selected (sentinel system) | Over 60% of samples sequenced | Weekly | Sequencing | https://lns.lu/departement/microbiologie/revilux/ |
| Latvia | Routine genomic surveillance | High availability | Positive RT-PCR samples | - | 150 or more samples sequenced per week | Weekly | Sequencing | 1)https://www.ecdc.europa.eu/en/publications-data/data-virus-variants-covid-19-eueea<br>2)https://eng.lsm.lv/article/society/health/colombian-strain-of-covid-19-detected-in-latvia.a408115/ |

| Country | Surveillance strategy | Sequencing availability | Target population | Sampling method | Sequenced volume | Reporting frequency | Diagnostic criteria | Source |
|---|---|---|---|---|---|---|---|---|
| Malta | Routine genomic surveillance | Moderate availability | Confirmed cases | - | At least 10% of samples sequenced | Weekly | Sequencing | 1)https://www.ecdc.europa.eu/en/publications-data/data-virus-variants-covid-19-eueea 2)https://newsbook.com.mt/en/covid-variants-malta-registers-a-total-of-442-cases-to-date/ 3)https://euobserver.com/coronavirus/151238 |
| Netherlands | Routine genomic surveillance | High availability | Positive PCR cases | Randomly selected | About 1000 samples sequenced per week | Weekly | Sequencing | https://www.rivm.nl/en/coronavirus-covid-19/research/pathogen-surveillance |
| Norway | Routine genomic surveillance | High availability | 1) Patients with covid-19 in hospital and ICU 2) Overseas traveler 3) Community patients 4) Vaccinated infected people | Sampling most in western Norway | 20-30% of samples sequenced | Weekly | Whole genome sequencing, PCR | https://www.fhi.no/en/publ/2020/weekly-reports-for-coronavirus-og-covid-19/ |
| Poland | Routine genomic surveillance | High availability | Confirmed cases | - | 150 or more samples sequenced per week; at least 5% of samples sequenced | Weekly | Sequencing | https://www.ecdc.europa.eu/en/publications-data/data-virus-variants-covid-19-eueea |
| Portugal | Routine genomic surveillance | High availability | Confirmed cases | - | 150 or more samples sequenced per week | Weekly | Sequencing | https://www.ecdc.europa.eu/en/publications-data/data-virus-variants-covid-19-eueea |
| Romania | Routine genomic surveillance | High availability | - | - | 150 or more samples sequenced per week | Weekly | Sequencing | https://www.ecdc.europa.eu/en/publications-data/data-virus-variants-covid-19-eueea |
| Russian Federation | Routine genomic surveillance since June 2020 | High availability | | | 7,000 samples sequenced since June 15, 2020 | Monthly | Sequencing | https://www.livemint.com/news/world/over-13-000-coronavirus-variants-detected-in-russia-vaccine-developer-11621673480754.html |
| Slovakia | Routine genomic surveillance | Moderate availability | Confirmed cases | - | 150 or more samples sequenced per week | Weekly | Sequencing | 1)https://www.ecdc.europa.eu/en/publications-data/data-virus-variants-covid-19-eueea |

| Country | Surveillance strategy | Sequencing availability | Target population | Sampling method | Sequenced volume | Reporting frequency | Diagnostic criteria | Source |
|---|---|---|---|---|---|---|---|---|
| | | | | | | | | 2)https://www.reuters.com/article/us-health-coronavirus-slovakia-idUKKBN2A52IJ<br>3) Brejová B et al. medRxiv 2021: 2021.07.13.21260431.[18]<br>4) https://euobserver.com/coronavirus/151238 |
| Slovenia | Routine genomic surveillance | Moderate availability | Confirmed cases | - | 150 or more samples sequenced per week; at least 5% of samples sequenced | Weekly | Sequencing | 1)https://www.ecdc.europa.eu/en/publications-data/data-virus-variants-covid-19-eueea<br>2)https://www.gov.si/en/news/2021-02-26-south-african-coronavirus-variant-now-in-slovenia/<br>3) https://euobserver.com/coronavirus/151238 |
| Sweden | Routine genomic surveillance | High availability | 1) COVID-19 patients<br>2) Individuals with suspected reinfection and breakthrough infection<br>3) Overseas traveler | Randomly selected (Representative) | 2000 or more samples sequenced per week; Over 10% of samples sequenced | Weekly | Whole genome sequencing, PCR | 1)https://www.folkhalsomyndigheten.se/smittskydd-beredskap/utbrott/aktuella-utbrott/covid-19/statistik-och-analyser/sars-cov-2-virusvarianter-av-sarskild-betydelse/<br>2)https://www.folkhalsomyndigheten.se/smittskydd-beredskap/utbrott/aktuella-utbrott/covid-19/statistik-och-analyser/sars-cov-2-virusvarianter-av-sarskild-betydelse/sars-cov-2-virusvarianter-av-sarskild-betydelse/instruktion-for-kompletterande-anmalan/ |
| Turkmenistan | | Moderate availability | | | | | | https://www.euro.who.int/en/countries/turkmenistan/news/news/2021/8/new-equipment-arrives-in-turkmenistan-to-strengthen-capacity-in-laboratory-diagnostics |

| Country | Surveillance strategy | Sequencing availability | Target population | Sampling method | Sequenced volume | Reporting frequency | Diagnostic criteria | Source |
|---|---|---|---|---|---|---|---|---|
| Bosnia and Herzegovina | | Moderate availability | COVID-19 positive cases | | | | whole genome sequencing | https://www.iaea.org/newscenter/news/bosnia-herzegovina-and-serbia-succeed-in-covid-19-virus-characterization-with-iaea/fao-support |
| Serbia | | Moderate availability | COVID-19 positive cases | | | | whole genome sequencing | https://www.iaea.org/newscenter/news/bosnia-herzegovina-and-serbia-succeed-in-covid-19-virus-characterization-with-iaea/fao-support |
| Turkey | | High availability | | | | | Sequencing | Karacan I et al. medRxiv 2020: 2020.12.25.20248851.[19] |
| **Region of Americas (AMR)** | | | | | | | | |
| Argentina | Routine genomic surveillance | High availability | 1) Positive international travelers;<br>2) Individuals with a positive result and no travel history;<br>3) Cases with a vaccination history | Representative sample of the different characteristics in 24 jurisdictions | 1261 samples were sequenced from Jan to Jun 7, 2021; 150 or more samples sequenced in recently week | Monthly, weekly | Whole and partial genome sequencing, PCR | 1)https://ianphi.org/_includes/documents/sections/news/2021/genomic-surveillance-perandones.pdf<br>2)http://pais.qb.fcen.uba.ar/files/reportes/pais-reporte20.pdf |
| Brazil | Routine genomic surveillance | High availability | 1) Imported cases<br>2) Cases that linked with area of circulation with B.1.1.7<br>3) Cases with epidemiological investigation | Sampled across the country (carried out by several labs) | 150 or more samples sequenced per week | Weekly | Sequencing | https://www.gov.br/saude/pt-br/media/pdf/2021/julho/02/69_boletim_epidemiologico_covid_2junho.pdf |
| Canada | Routine genomic surveillance | High availability | 1) International travelers and close contacts<br>2) Individuals with S-gene failure<br>3) Suspected reinfection individuals | Randomly selected | 1000 or more samples sequenced per day | Daily | Sequencing, PCR (only in Ontario) | 1) Genome Canada Canadian COVID-19 Genomics Network (CanCOGeN) et al. Can Commun Dis Rep 2021; 47(3): 139-41.[20] |

| Country | Surveillance strategy | Sequencing availability | Target population | Sampling method | Sequenced volume | Reporting frequency | Diagnostic criteria | Source |
|---|---|---|---|---|---|---|---|---|
| | | | 4) Severe cases younger than 50 years old without significant comorbidities<br>5) Vaccinated individuals with subsequent SARS-CoV-2 infection<br>6) Known or suspected super spreading events | | | | | 2） https://www.publichealthontario.ca/en/laboratory-services/test-information-index/covid-19-voc<br><br>3)https://www.ctvnews.ca/health/coronavirus/tracking-variants-of-the-novel-coronavirus-in-canada-1.5296141 |
| United States | Routine genomic surveillance | High availability | RT-PCR positive confirmed cases with Ct value ≤ 28 | Randomly selected | 10,000 or more samples sequenced per week; less than 5% of samples sequenced | Weekly or bi-weekly | Sequencing | 1) https://www.cdc.gov/coronavirus/2019-ncov/variants/variant-surveillance.html<br>2)https://www.aphl.org/programs/preparedness/Crisis-Management/Documents/2021.04.09_NS3_REVISED.pdf |
| Uruguay | Routine genomic surveillance | High availability | SARS-CoV-2–positive samples in Uruguay | | 100-300 samples sequenced per week | Weekly | PCR, Sequencing | Rego N, et al. Emerging Infectious Disease 2021; 27(11).[21] |
| Peru | Routine genomic surveillance | High availability | 1) Local representative positive samples<br>2) Imported cases<br>3) Outbreak-based cases | Representative samples | 1200 samples sequenced per month | | Sequencing | https://web.ins.gob.pe/es/prensa/noticia/la-guerra-silenciosa-del-instituto-nacional-de-salud |
| Chile, Colombia, Mexico, Panama, El Salvador, Trinidad and Tobago | - | High availability | | | | | Whole genome sequencing (In-country) | 1.https://www.paho.org/en/topics/influenza/covid-19-genomic-surveillance-regional-network<br>2.https://www.paho.org/en/news/21-7-2021-regional-genomic-surveillance-network-tracks-covid-19-virus-variants-throughout |

| Country | Surveillance strategy | Sequencing availability | Target population | Sampling method | Sequenced volume | Reporting frequency | Diagnostic criteria | Source |
|---|---|---|---|---|---|---|---|---|
| Costa Rica | Routine genomic surveillance | High availability | positive samples with Ct value ≤ 25 or being symptomatic | Representative samples when possible | laboratories must send up to 10 positive samples per week | | Sequencing | 1.https://www.ministeriodesalud.go.cr/index.php/centro-de-prensa/noticias/741-noticias-2020/1642-inciensa-logra-secuenciar-el-genoma-completo-del-nuevo-coronavirus-sars-cov-2-covid-19 2.https://www.ministeriodesalud.go.cr/sobre_ministerio/prensa/docs/ls_vs_001_version_19_vigilancia_covid19_31032021.pdf 3. Molina-Mora JA, et al. Infect Genet Evol.2021;92:104872.[22] |
| The Bahamas, Bolivia, Barbados, Dominican Republic, Guatemala, Guyana, Honduras, Haiti, Jamaica, Paraguay, Suriname, Venezuela | - | Moderate availability | | | | | Whole genome sequencing (Sending-out for external sequencing) | https://www.paho.org/en/topics/influenza/covid-19-genomic-surveillance-regional-network |
| Ecuador | No routine genomic surveillance | Moderate availability | | | 119 samples sequenced from Mar 2020 to Jan 2021 | | Sequencing (MinION platform) | Márquez S, et al. medRxiv 2021: 2021.03.19.21253620.[23] |

| Country | Surveillance strategy | Sequencing availability | Target population | Sampling method | Sequenced volume | Reporting frequency | Diagnostic criteria | Source |
|---|---|---|---|---|---|---|---|---|
| Cuba | Limited routine genomic surveillance | High availability | serious and critical cases, deaths, travelers and in outbreaks of the disease | | 1064 samples sequenced from Jan to July, 2021 | | Sequencing | 1)https://salud.msp.gob.cu/variantes-de-sars-cov-2-en-cuba-motivo-mas-para-fortalecer-las-medidas-de-aislamiento/ 2)https://salud.msp.gob.cu/variantes-geneticas-aumentan-la-severidad-de-la-covid-19/ |
| Trinidad and Tobago | - | Moderate availability | | | | | Ship samples to University of the West Indies for sequencing | https://health.gov.tt/brazilian-variant-found-in-additional-covid-19-samples |
| Saint Kitts and Nevis | - | Moderate availability | | | | | PCR | https://iris.paho.org/bitstream/handle/10665.2/54588/COVID-19SitRep55_eng.pdf?sequence=1&isAllowed=y |
| Saint Lucia | No routine genomic surveillance | Moderate availability | | | Less than ten samples sequenced per week | | | https://www.covid19response.lc/blogs/saint-lucia-confirms-sars-cov-2-british-variant-in-country |
| Antigua and Barbuda | No routine genomic surveillance | Moderate availability | | | Eight samples taken between July 18 and 22, 2021 | | sequencing | 1.https://ab.gov.ag/detail_page.php?page=2.php 2.https://ab.gov.ag/media_page.php?page=425 3.https://ab.gov.ag/pdf/Covid_19_Variant_Update.pdf |
| **South-East Asia Region (SEAR)** | | | | | | | | |
| Bangladesh | Limited routine genomic surveillance | High availability | RT-PCR positive cases with Ct value < 30 | Randomly selected | 748 samples sequenced from Dec 2020 to Jun 2021 | Weekly | Whole-genome sequencing | 1.https://ianphi.org/_includes/documents/sections/news/2021/experience-of-iedcr-in-covid-19-vaccination-shirin.pdf 2.https://old.iedcr.gov.bd/website/images/files/nCoV/FourthJulyCOVID19Update.pdf |

| Country | Surveillance strategy | Sequencing availability | Target population | Sampling method | Sequenced volume | Reporting frequency | Diagnostic criteria | Source |
|---|---|---|---|---|---|---|---|---|
| | | | | | | | | 3. https://www.icddrb.org/news-and-events/news?id=883&task=view<br>4. Hossain ME, et al. Microbiol Resour Announc 2021; 10(8).[24]<br>5. Saha S, et al. BMJ Glob Health 2021; 6(5).[25] |
| Indonesia | Routine genomic surveillance | High availability | 1) Specimens from imported travelers testing positive<br>2) Vaccine failures<br>3) Reinfections<br>4) Severe illness and death<br>5) Outbreaks with unusual characteristics | - | 150 or more samples sequenced per week | Weekly | Whole genome sequencing | 1.https://www.litbang.kemkes.go.id/assets/2021/06/Update_VoC-COVID-19-20-juni-2021.pdf<br>2.https://drive.google.com/drive/folders/1ocT30cqXeDsrr72YJA-bdTzxudolaOZT<br>3.https://www.litbang.kemkes.go.id/short-report-of-working-group-surveillance-of-covid-19/ |
| India | Routine genomic surveillance | High availability (28 National Laboratories) | 1) RT-PCR positive cases with Ct value < 30<br>2) Vaccinated positive samples<br>3) Post-infected and re-infected positive samples | Randomly selected | 150 or more samples sequenced per week; 5% of samples sequenced | - | Whole genome sequencing | 1.https://www.mohfw.gov.in/pdf/IndianSARSCoV2PDFGenomicsConsortiumGuidanceDocument.pdf<br>2.http://clingen.igib.res.in/covid19genomes/<br>3.https://dbtindia.gov.in/insacog |
| Sri Lanka | Limited routine genomic surveillance | Moderate availability | RT-PCR positive cases with Ct value < 30 | Several rounds of sequencing had been performed | 15 or more samples sequenced per week | - | Sequencing | 1.http://www.health.gov.lk/moh_final/english/public/elfinder/files/publications/2021/Edited%20Final%20SPRP%20on%2015th%20May%202021%20.pdf<br>2. Jeewandara C, et al. medRxiv 2021: 2021.05.05.21256384.[26] |

| Country | Surveillance strategy | Sequencing availability | Target population | Sampling method | Sequenced volume | Reporting frequency | Diagnostic criteria | Source |
|---|---|---|---|---|---|---|---|---|
| Thailand | Routine genomic surveillance | High availability | RT-PCR positive cases | Surveillance network of laboratories across the country | 150 or more samples sequenced per week | Weekly | Sequencing, PCR | https://www3.dmsc.moph.go.th/post-group/10 |
| Bhutan | - | Moderate availability | | | | | Sequencing (Send to Thailand) | https://kuenselonline.com/bhutan-saw-seven-variants-of-sars-cov-2-virus/ |
| Nepal | | Moderate availability | SARS-CoV-2 positive samples | | | | Sequencing | https://cdn.who.int/media/docs/default-source/nepal-documents/novel-coronavirus/who-nepal-sitrep/weekly-who-nepal-situation-updates_66_v2.pdf?sfvrsn=1197fcb0_5 |
| Timor-Leste | No routine genomic surveillance | Moderate availability | | | 70 positive samples sequenced between May 2020 and March 2021 | | Sequencing | http://www.tatoli.tl/en/2021/04/14/timor-leste-reports-17-new-local-cases-of-covid-19-and-14-recoveries/ |
| Maldives | | Moderate availability | | | | | Sequencing | https://raajje.mv/101870 |
| **Western Pacific Region (WPR)** | | | | | | | | |
| Australia | Routine genomic surveillance | High availability | All suitable available samples excluding too low viral loads | Nearly all | 61% of positive cases sequenced (as of 18 July) | Weekly | Sequencing | 1) https://www1.health.gov.au/internet/main/publishing.nsf/Content/1D03BCB527F40C8BCA258503000302EB/$File/covid_19_australia_epidemiology_report_36_reporting_period_ending_28_february_2021.pdf<br>2) https://www.cdgn.org.au/austrakka<br>3)https://www1.health.gov.au/internet/main/publishing.nsf/Content/1D03BCB527F40C8BCA258503000302EB/$File/covid_19_australia_epidemiology_reporting_last_updated_11_may_2021.pdf |

| **Country** | **Surveillance strategy** | **Sequencing availability** | **Target population** | **Sampling method** | **Sequenced volume** | **Reporting frequency** | **Diagnostic criteria** | **Source** |
|---|---|---|---|---|---|---|---|---|
| China | Routine genomic surveillance | High availability | Confirmed imported cases with enough virus load | Nearly all since Sep 2020) | Nearly all | Weekly | Sequencing | Internal data from China CDC |
| Japan | Routine genomic surveillance | High availability | 1) People who have traveled abroad in the last two weeks<br>2) Domestic cases | - | 62,861 samples sequenced as of Aug 16, 2021 | | Sequencing and PCR | 1) https://www.niid.go.jp/niid/ja/diseases/ka/corona-virus/2019-ncov/2484-idsc/10554-covid19-52.html<br>2)https://www.niid.go.jp/niid/ja/diseases/ka/corona-virus/2019-ncov/10220-covid19-36.html |
| South Korea | Routine genomic surveillance | High availability | Positive samples of confirmed cases related to various domestic outbreaks and imported cases | Representative samples | 14.7% of samples sequenced in April 2021 | | Partial and whole genome Sequencing | 1.http://www.kdca.go.kr/board/board.es?mid=a30501000000&bid=0031&list_no=713847&act=view<br>2. Park AK, et al. Osong public health and research perspectives 2021; 12(1): 37-43.[27] |
| Laos | No routine genomic surveillance | High availability | 1) Local cases<br>2) Imported cases | | 37 samples sequenced from 4 Aug to 17 Aug | Bi-weekly | Sequencing | https://www.who.int/laos/emergencies/covid-19-in-lao-pdr/situation-reports |
| Malaysia | Limited routine genomic surveillance | High availability | 1) Individuals who had been to UK since Oct 2020 and positive cases with a travel history to other countries<br>2) RT-PCR positive cases which met the cut off for Ct value or from local clusters<br>3) Individuals take on the sequencing works<br>4) Cases of death due to COVID-19 | - | 15 or more samples sequenced per week | Irregularly | Whole genome sequencing | 1) http://covid-19.moh.gov.my/semasa-kkm/2021/05/mutasi-spike-protein-di-malaysia-17052021<br>2) http://covid-19.moh.gov.my/semasa-kkm/2021/01/uk-traveller-samples-since-october2020<br>3) http://covid-19.moh.gov.my/semasa-kkm/2021/04/maklumat-terkini-mutasi-protein-spike-di-malaysia |

| Country | Surveillance strategy | Sequencing availability | Target population | Sampling method | Sequenced volume | Reporting frequency | Diagnostic criteria | Source |
|---|---|---|---|---|---|---|---|---|
| New Zealand | Routine genomic surveillance | High availability | Confirmed cases | | At least 10% of samples sequenced | Weekly | Real-time whole genome sequencing | 1.https://www.health.govt.nz/system/files/documents/pages/6_july_2021_-_variants_update_-_full_report.pdf<br>2. Douglas J, et al. Emerging Infectious Disease 2021; 27(9).[28] |
| Philippines | Routine genomic surveillance | High availability | 1) local cases<br>2) Returning Overseas Filipinos | | 150 or more samples sequenced per week | Irregularly | Whole genome sequencing | 1.https://doh.gov.ph/press-release/DOH-UP-PGC-AND-UP-NIH-DETECT-MORE-COVID-19-VARIANTS-IN-ONGOING-BIOSURVEILLANCE<br>2.https://doh.gov.ph/doh-press-release/BIOSURVEILLANCE-DETECTS-SARS-COV-2-MUTATIONS-FURTHER-INVESTIGATION-NEEDED-TO-DETERMINE-PUBLIC-HEALTH-IMPLICATION |
| Singapore | Routine genomic surveillance | High availability | All COVID-19 cases | All COVID-19 cases | All COVID-19 cases sequenced | | Whole genome sequencing | https://www.moh.gov.sg/news-highlights/details/investigations-into-13-covid-19-cases-who-served-shn-at-mandarin-orchard-singapore |
| Fiji | - | Moderate availability | | | | | Sequencing | https://www.rnz.co.nz/international/pacific-news/441313/covid-19-cases-continue-to-climb-in-fiji |
| Cambodia | Routine genomic surveillance | High availability | 1) SARS-CoV-2 specimens from land borders<br>2) Community-acquired samples | | By 16 August 2021, a total of 2,865 samples have been tested | Weekly | PCR, whole genome sequencing | https://www.who.int/cambodia/internal-publications-detail |

| Country | Surveillance strategy | Sequencing availability | Target population | Sampling method | Sequenced volume | Reporting frequency | Diagnostic criteria | Source |
|---|---|---|---|---|---|---|---|---|
| | | | 3) Returning migrants. | | | | | |
| Papua New Guinea | - | Moderate availability | | | | | | https://www.facebook.com/PNGNDOH/posts/4117440295005664?__tn__=K-R |
| Vietnam | - | High availability | 1) High-risk groups<br>2) COVID-19 patients | | | | Sequencing | 1. http://news.chinhphu.vn/Home/India-and-UK-variants-behind-new-COVID19-cases-in-Viet-Nam/20215/43665.vgp<br>2. Nguyen Van Vinh Chau et al. J Infect. 2021; 82(6):276-316. [29] |
| Mongolia | - | Moderate availability | | | | | | 1.https://montsame.mn/en/read/258007<br>2.https://www.who.int/mongolia/news/detail/09-04-2021-australia-and-world-health-organization-work-together-to-increase-covid-19-testing-capacity-in-mongolia |
| Brunei | Samples have been sent to a laboratory in Singapore for genomic sequencing | Moderate availability | | | | | sequencing | https://www.who.int/brunei/internal-publications-detail |
| **African Region (AFR)** | | | | | | | | |
| Kenya, Senegal, Uganda | Routine genomic surveillance | High availability | | | | Weekly | Sequencing | 1.https://www.arcgis.com/home/webmap/viewer.html?url=https://services8.arcgis.com/vWozsma9VzGndzx7/ArcGIS/rest/services/Test_Africa_PGI_DB_GR/FeatureServer/0&source=sd (Source for all African countries)<br>2.https://www.afro.who.int/publications/interim-operational-guidance-sars-cov-2-genomic- |

| Country | Surveillance strategy | Sequencing availability | Target population | Sampling method | Sequenced volume | Reporting frequency | Diagnostic criteria | Source |
|---|---|---|---|---|---|---|---|---|
| | | | | | | | | surveillance-africa-updated-guide (Source for all African countries) |
| Angola, The Gambia, Mozambique, Zimbabwe | Routine genomic surveillance | Moderate availability | | | | Weekly | Sequencing | Same as above |
| Benin, Burkina Faso, Botswana, Comoros, Cameroon, Cape Verde, Ethiopia, Guinea, Lesotho, Madagascar, Malawi, Namibia, Sierra Leone, Swaziland. Togo | Limited routine genomic surveillance | Moderate availability | | | | Weekly | Sequencing | Same as above |
| Gabon | No routine genomic surveillance | High availability | | | | | | Same as above |
| Mauritius | Routine genomic surveillance | Moderate availability | | | 124 of 432 positive cases sequenced in March 2021 | | Sequencing | Tegally H, et al. medRxiv 2021: 2021.06.16.21259017.[30] |
| Mali | - | Moderate availability | | | | | | Dara A, et al. bioRxiv 2021: 2021.05.05.442742.[31] |
| Rwanda | Limited routine genomic surveillance | Moderate availability | | | 1.2 % of samples sequenced | | | 1. Yvan Butera EM, et al. medRxiv 2021: 2021.04.02.21254839.[32] |
| Côte d'Ivoire | Limited routine genomic surveillance | Moderate availability | RT-PCR positive samples | Randomly selected | | Weekly | Sequencing | 1. Wilkinson E, et al. medRxiv 2021: 2021.05.12.21257080.[33] |
| Democratic Republic of Congo (DRC) | Limited routine genomic surveillance | High availability | 1) Imported cases<br>2) RT-PCR positive samples | Randomly selected | 1.4 % of samples sequenced | Weekly | Sequencing | 1. Wilkinson E, et al. medRxiv 2021: 2021.05.12.21257080.[33] |

| Country | Surveillance strategy | Sequencing availability | Target population | Sampling method | Sequenced volume | Reporting frequency | Diagnostic criteria | Source |
|---|---|---|---|---|---|---|---|---|
| Congo | Limited routine genomic surveillance | Moderate availability | RT-PCR positive samples with a Ct values <30 | Randomly selected | | Weekly | Next-generation sequencing | 1. Ntoumi F, et al. Int J Infect Dis 2021: 105: 735-8.[34] |
| Algeria | No routine genomic surveillance | Moderate availability | 1) Imported cases<br>2) RT-PCR positive samples<br>3) Cluster/outbreak investigations | Randomly selected | 0.08% of samples sequenced | Weekly | PCR, Sanger Sequencing, whole genome sequencing | 1. Wilkinson E, et al. medRxiv 2021: 2021.05.12.21257080.[33] |
| Ghana (Uhas) | Routine genomic surveillance | High availability | 1) Cluster/outbreak investigations<br>2) Confirmed cases in periods of suspected widespread infections | Randomly selected | | Weekly | Sequencing | 1. Wilkinson E, et al. medRxiv 2021: 2021.05.12.21257080.[33] |
| Equatorial Guinea | Routine genomic surveillance | Moderate availability | 1) Imported cases<br>2) RT-PCR positive samples<br>3) Cluster/outbreak investigations | Randomly selected | 3.10% | Weekly | Sequencing | 1. Wilkinson E, et al. medRxiv 2021: 2021.05.12.21257080.[33] |
| Nigeria | Routine genomic surveillance | High availability | | Randomly selected | | Weekly | Sequencing | 1.https://covid19.ncdc.gov.ng/media/files/STATEMENT_ON_VARIANTS_OF_SARS-COV-2_IN_NIGERIA_ZyRAI48.pdf |
| South Africa | Routine genomic surveillance | High availability | 1) Routine specimens with representative of multiple geographic regions<br>2) Special interest specimens, such as vaccine breakthrough cases, re-infection cases, etc. | Randomly selected | Approximately 10%-20% of randomly selected positive samples | Bi-weekly | Next-generation sequencing | 1.https://www.krisp.org.za/manuscripts/Update_SA_sequencing_1_July_2021_AvG.pdf |
| Zambia | Routine genomic surveillance | Moderate availability | 1) Imported cases<br>2) RT-PCR positive samples<br>3) Cluster/outbreak investigations<br>4) Re-infection cases | Randomly selected | | Weekly | Sequencing | 1. Wilkinson E, et al. medRxiv 2021: 2021.05.12.21257080.[33] |

| Country | Surveillance strategy | Sequencing availability | Target population | Sampling method | Sequenced volume | Reporting frequency | Diagnostic criteria | Source |
|---|---|---|---|---|---|---|---|---|
| Other African countries | | Moderate availability | | | | | | https://www.afro.who.int/publications/interim-operational-guidance-sars-cov-2-genomic-surveillance-africa-updated-guide |
| **Eastern Mediterranean Region (EMR)** | | | | | | | | |
| Afghanistan, Bahrain, Djibouti, Iraq, Sudan, Somalia, Syria, Yemen | No routine genomic surveillance* | Low availability | | | | | | http://www.emro.who.int/health-topics/corona-virus/situation-reports.html (Source for all EMR countries) |
| United Arab Emirates, Iran, Jordan, Kuwait, Oman, Saudi Arabia, Tunisia | - | High availability | | | | | | Same as above |
| Pakistan | | Moderate availability | | | | | PCR, genome sequencing | http://www.emro.who.int/pak/pakistan-news/who-and-national-institute-of-health-collaborate-to-detect-sars-cov-2-variants-of-interest-and-concern-through-pcr-detection-and-genome-sequencing.html |
| Lebanon | Limited routine genomic surveillance | High availability | Clinical diagnostic samples | Randomly selected | | | High throughput genome sequencing | Merhi G, et al. medRxiv 2021:2021.08.10.21261847.[35] |
| Morocco, | Routine genomic surveillance | High availability | 1) Cluster/outbreak investigations<br>2) Imported cases<br>3) Re-infection cases | | Sequencing of 10% of sample that are positive for S drop real time PCR test | | Sequencing, PCR | Wilkinson E, et al. medRxiv 2021: 2021.05.12.21257080.[33] |

| Country | Surveillance strategy | Sequencing availability | Target population | Sampling method | Sequenced volume | Reporting frequency | Diagnostic criteria | Source |
|---|---|---|---|---|---|---|---|---|
| Qatar | Limited routine genomic surveillance | High availability | Clusters cases | Samples selection was based on epidemiological characteristics | 1.5% of samples sequenced from Mar 2020 to Mar 2021 | | Whole-genome sequencing | Benslimane FM, et al. medRxiv 2021: 2021.05.19.21257433.[36] |
| Egypt | Routine genomic surveillance | High availability | | | | Weekly | Sequencing | https://www.arcgis.com/home/webmap/viewer.html?url=https://services8.arcgis.com/vWozsma9VzGndzx7/ArcGIS/rest/services/Test_Africa_PGI_DB_GR/FeatureServer/0&source=sd |
| Libya | Limited routine genomic surveillance | Moderate availability | | | | Weekly | Sequencing | 1.https://www.arcgis.com/home/webmap/viewer.html?url=https://services8.arcgis.com/vWozsma9VzGndzx7/ArcGIS/rest/services/Test_Africa_PGI_DB_GR/FeatureServer/0&source=sd 2.https://www.afro.who.int/publications/interim-operational-guidance-sars-cov-2-genomic-surveillance-africa-updated-guide |

* For countries where genomic surveillance information was unavailable, we assumed that countries with low sequencing availability didn’t carry out routine genomic surveillance.

**Table S5. Completeness analysis of metadata collected from GISAID dataset, as of Aug 20, 2021.**

| WHO region | Total (N) | National geography [a] | Subnational geography [a] | Age | Sex | Date of collection | Patient status [b] | Vaccinated status [c] | Lineage |
|---|---|---|---|---|---|---|---|---|---|
| AFR | 31254 | 100.0% | 91.6% | 88.0% | 89.7% | 97.3% | 14.4% | 0.4% | 96.5% |
| AMR | 1048063 | 100.0% | 99.7% | 54.2% | 55.3% | 96.2% | 3.3% | 0.2% | 98.8% |
| EMR | 11868 | 100.0% | 76.9% | 64.7% | 65.4% | 94.6% | 16.7% | 0.2% | 93.1% |
| EUR | 1827918 | 100.0% | 91.9% | 22.2% | 20.8% | 97.1% | 2.9% | 0.1% | 97.2% |
| SEAR | 60085 | 100.0% | 98.7% | 82.9% | 83.5% | 93.8% | 10.3% | 1.6% | 96.9% |
| WPR | 137009 | 100.0% | 52.0% | 33.5% | 34.4% | 96.2% | 13.4% | 0.0% | 96.8% |
| Global | 3116197 | 100.0% | 92.8% | 35.5% | 35.1% | 96.7% | 3.8% | 0.1% | 97.7% |

[a] The completeness of geographic information (national or subnational) reported in metadata file.

[b] The completeness of information about patient status, includes the symptomatic history, clinical severity or outcome, etc.

**c** The completeness of vaccinated status, includes whether vaccinated or not, vaccinated platform, vaccinated dose, or vaccinated time, etc.

**Abbreviation**: AFR, African Region; AMR, Region of Americas; EMR, Eastern Mediterranean Region; EUR, European Region; SEAR, South-East Asia Region; WPR, Western Pacific Region

**Table S6. The cumulative official number of variants that used for calculating the public availability extent of genomic data.**

| Country | ISO week in 2021 [a] | Alpha [b] | Beta | Gamma | Delta |
|---|---|---|---|---|---|
| African Region | | | | | |
| Algeria | 31 | 8 | - [c] | - | - |
| South Africa | 31 | 215 | 6769 | - | 5046 |
| Democratic Republic of the Congo | 31 | 16 | 32 | - | 228 |
| Kenya | 31 | 562 | 191 | - | 575 |
| Malawi | 31 | 5 | 333 | - | 121 |
| The Gambia | 31 | 76 | - | - | 154 |
| Senegal | 31 | 219 | 3 | - | 264 |
| Region of Americas | | | | | |
| Canada | 31 | 229356 | 2399 | 21331 | 33547 |
| Argentina | 33 | 353 | 1 | 2212 | 235 |
| Peru | 31 | 19 | - | 998 | 192 |
| Eastern Mediterranean Region | | | | | |
| United Arab Emirates | 11 | 25 | 5 | 2 | - |
| Jordan | 11 | 3389 | 5 | 4 | - |
| Libya | 11 | 25 | 15 | - | - |
| Iran | 11 | 1792 | - | - | - |
| Morocco | 11 | 115 | - | - | - |
| Tunisia | 11 | 192 | - | - | - |
| Iraq | 11 | 236 | - | - | - |
| Afghanistan | 11 | 7 | - | - | - |
| Saudi Arabia | 11 | 10 | - | - | - |
| European Region | | | | | |
| Denmark | 33 | 68359 | 131 | 66 | 31314 |
| Finland | 28 | 7953 | 1445 | 7 | 2876 |
| Austria | 33 | 132033 | 1351 | 153 | 19859 |
| Sweden | 33 | 71612 | 2681 | 206 | 10819 |
| Spain | 29 | 22415 | 1320 | 957 | 10369 |
| Luxembourg | 32 | 6971 | 1196 | 1215 | 1555 |
| Netherlands | 29 | 26014 | 439 | 374 | 11072 |
| Ireland | 28 | 15954 | 77 | 30 | 4245 |
| Belgium | 30 | 23164 | 1441 | 2196 | 12797 |

| | | | | | |
|---|---|---|---|---|---|
| Cyprus | 27 | 666 | 1 | - | 299 |
| Estonia | 30 | 5275 | 51 | 9 | 1725 |
| France | 30 | - | 3175 | 1645 | 62122 |
| Greece | 30 | 16294 | 545 | 12 | 7325 |
| Hungary | 30 | 700 | - | 1 | 655 |
| Iceland | 30 | 535 | 4 | 16 | 3491 |
| Italy | 30 | 27755 | 286 | 2852 | 13533 |
| Latvia | 30 | 3570 | 14 | 4 | 1247 |
| Lithuania | 29 | 9685 | 12 | 6 | 1695 |
| Norway | 30 | 39265 | 625 | 11 | 11717 |
| Poland | 30 | 19751 | 65 | 26 | 965 |
| Portugal | 30 | 13771 | 112 | 184 | 5818 |
| Romania | 30 | 1541 | 8 | 20 | 598 |
| Slovakia | 30 | 9377 | 28 | - | 878 |
| United Kingdom | 33 | 277634 | 1093 | 272 | 535387 |
| South-East Asia Region | | | | | |
| India | 31 | 4935 | 246 | 4 | 32617 |
| Thailand | 32 | 14222 | 545 | - | 15973 |
| Indonesia | 31 | 64 | 17 | - | 2229 |
| Western Pacific Region | | | | | |
| Australia | 31 | 554 | 99 | 8 | 7735 |
| Philippines | 33 | 2395 | 2669 | - | 1789 |
| Malaysia | 28 | - | 209 | - | 424 |
| New Zealand | 33 | 186 | 33 | 8 | 245 |
| Japan | 33 | 43436 | 116 | 121 | 12509 |
| Cambodia | 33 | - | - | - | 999 |
| China | 20 | 102 | 46 | 3 | 38 |
| South Korea | 16 | 551 | 71 | 10 | - |
| Laos | 29 | 63 | - | - | 62 |

[a] A fixed three-week delay was used to extrapolate the date of specimen collection for Canada, Peru, Finland, Spain, Netherlands, Ireland, Belgium, Cyprus, Estonia, France, Greece, Hungary, Iceland, Italy, Latvia, Lithuania, Norway, Poland, Portugal, Romania, Slovakia, Australia, Malaysia, China, Laos, India, Indonesia, all countries in Africa and Eastern Mediterranean Region.

[b] As of one ISO week, the cumulative official number of Alpha variants in one country.

[c] No cases has been detected or the data was unavailable.

### Supplementary Figures

#### Fig S1. Distribution of 194 Member States by WHO region.

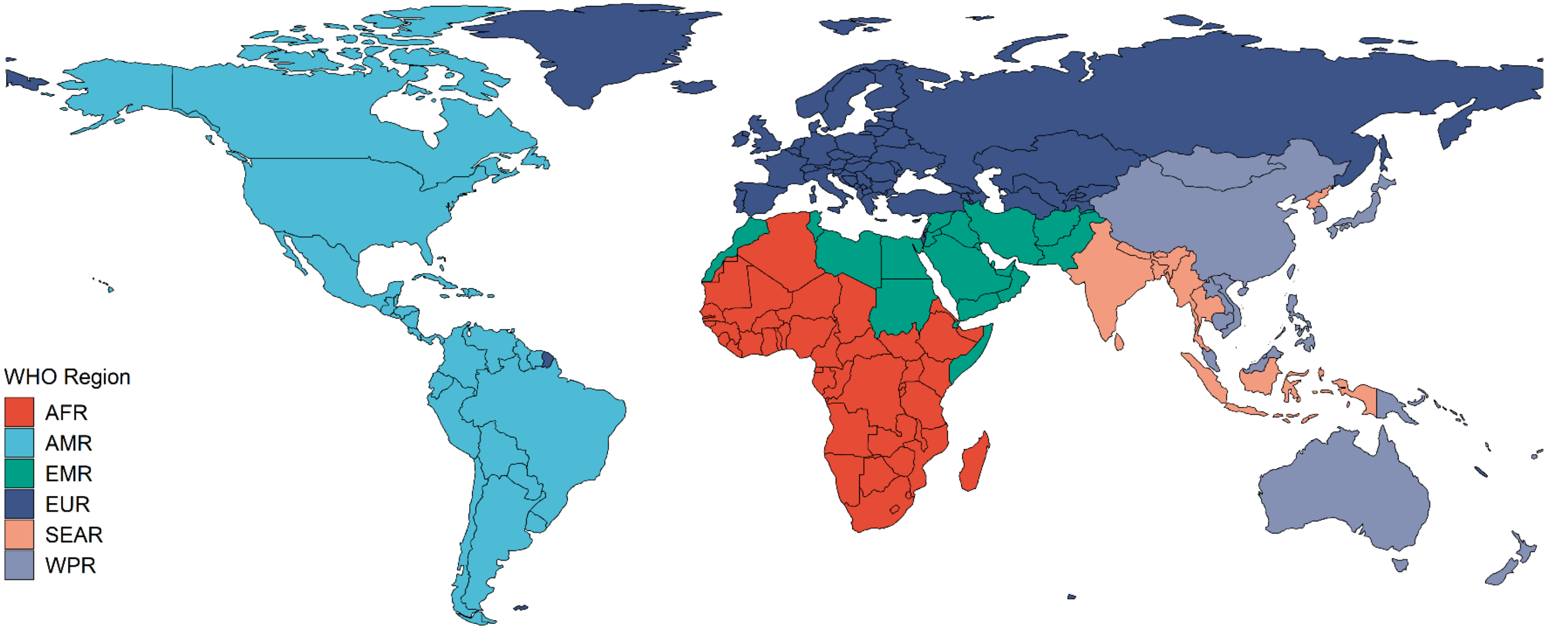

**Fig S2. Country classification based on the identification of variants of concern (VOCs) and the sharing status of sequence.**

The information about whether identified VOC or not in each country was derived from WHO situation report[37]. Data are as of 20 August 2021. NA, data not applicable.

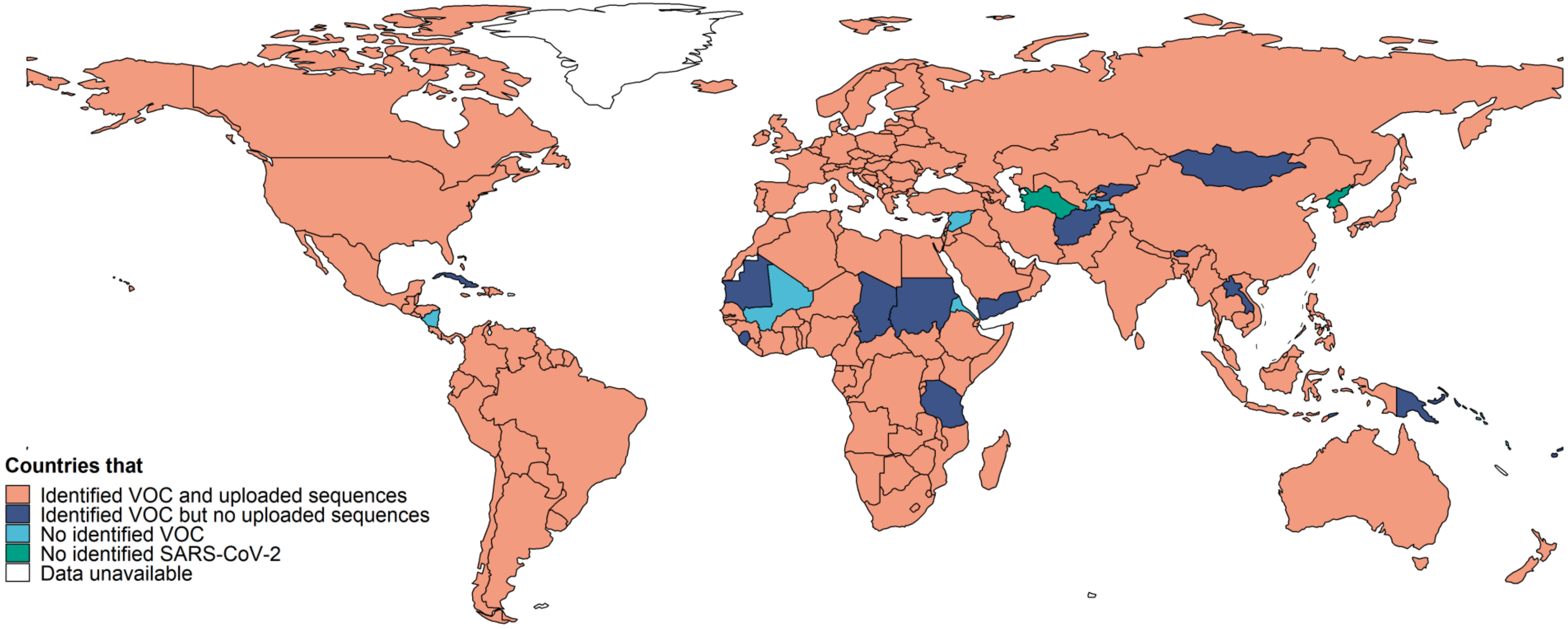

**Fig S3. The dynamic prevalence of variants of interest (VOIs) by time and country.**

As of 2 September 2021, WHO has designated Eta, Iota, Kappa, Lambda and Mu as VOIs.

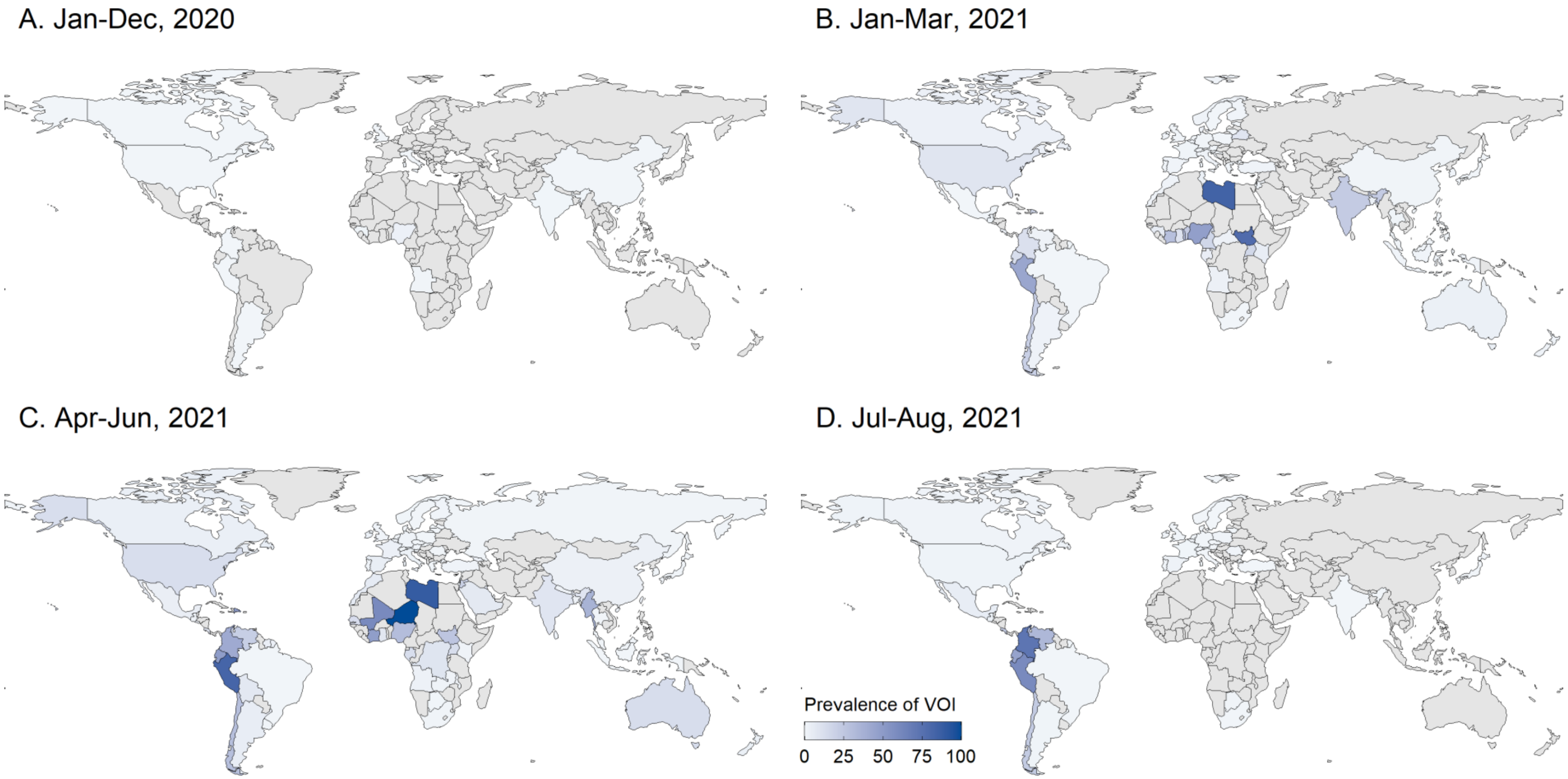
